## Supplementary Figure S1 for "Long-range PCR-based NGS applications to diagnose Mendelian retinal diseases" (from another version of this preprint: version 1, 2020-12-02)

**Figure S1. LR PCRs panel coverage results.**

Screenshot from the Alamut Visual software showing alignment of WES assay compared to the LR PCR results for the panel loci (in the same order reported on Table 1).

The loci are *PPT1* (A), *CRB1* (B), *PCARE* (C), *EFEMP1* (D), *IMPG2* (E), *PROM1* (F), *MFSD8* (G), *CTNNA1* (H), *GUCA1A* (I), *GUCA1B* (J), *PRPH2* (K), *IMPG1* (L), *ELOVL4* (M), *RP1L1* (N), *RP1* (O), *CNGB3* (P), *KCNV2* (Q), *ATOH7* (R), *PDE6C* (S), *BEST1* (T), *C1QTNF5* (U), *PDE6H* (V), *RDH5* (W), *OTX2* (X), *NR2E3* (Y), *RLBP1* (Z), *GUCY2D* (AA), *FSCN2* (BB), *RAX2* (CC), *TIMP3* (DD), *RS1* (EE), *RPGR* (FF), and *RP2* (GG). Since *DHS6S1* is not a protein-coding locus, it is not covered by Alamut Visual. Therefore, coverage and alignment are shown on IGV (HH).

A)

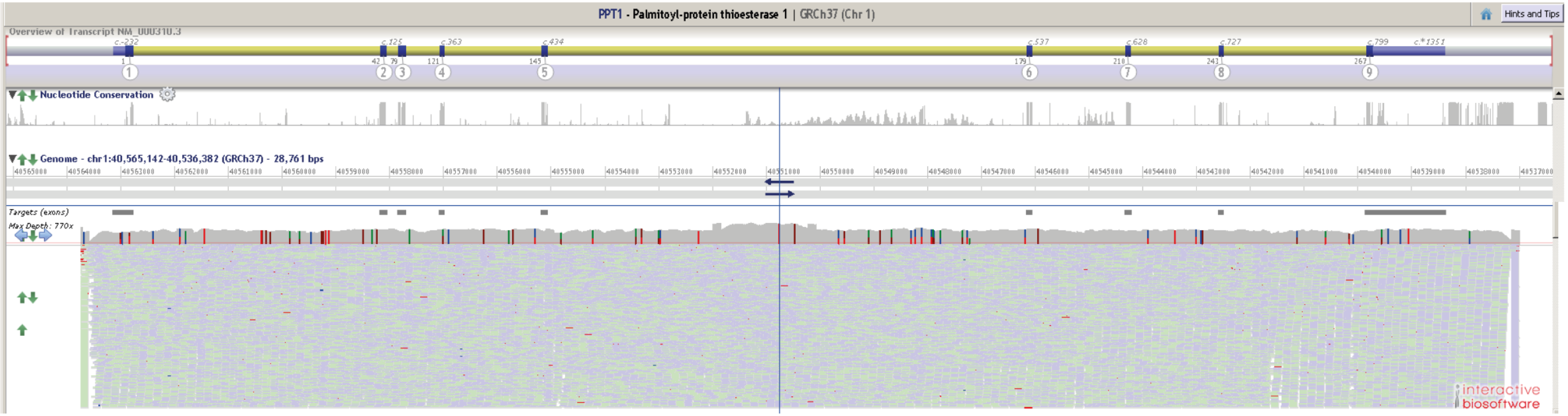

B)

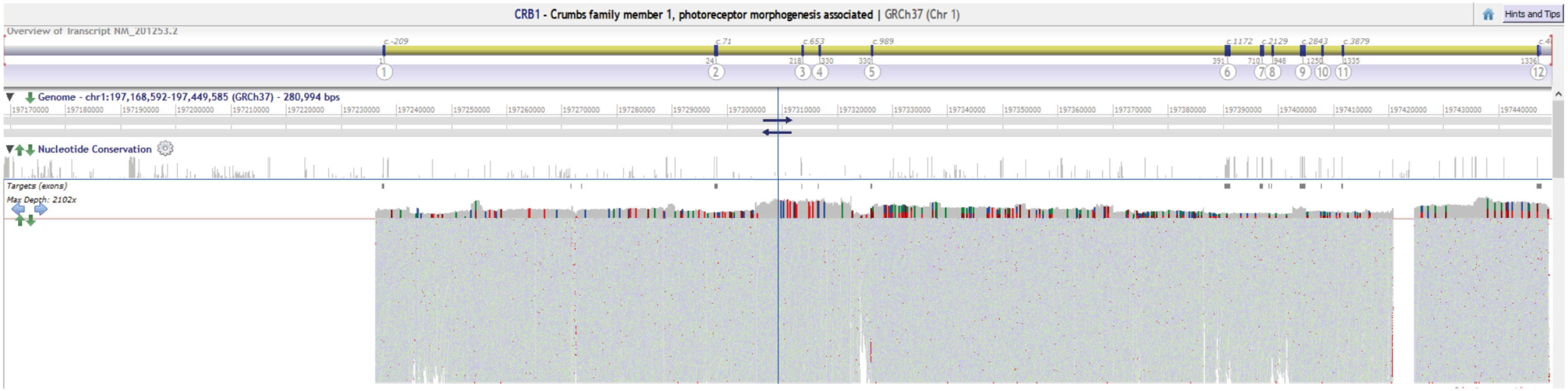
CRB1 - Crumbs family member 1, photoreceptor morphogenesis associated | GRCh37 (Chr 1)
Hints and Tips
Overview of Transcript NM_201253.2
Genome - chr1:197,168,592-197,449,585 (GRCh37) - 280,994 bps
Nucleotide Conservation
Targets (exons)
Max Depth: 2102x

C)

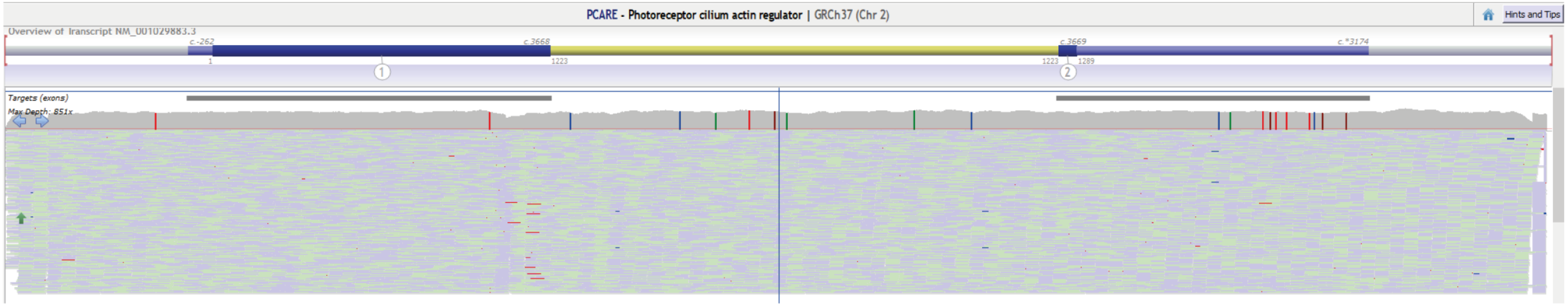
PCARE - Photoreceptor cilium actin regulator | GRCh37 (Chr 2)
Hints and Tips
Overview of Transcript NM_001029883.3
Targets (exons)
Max Depth: 851x

D)

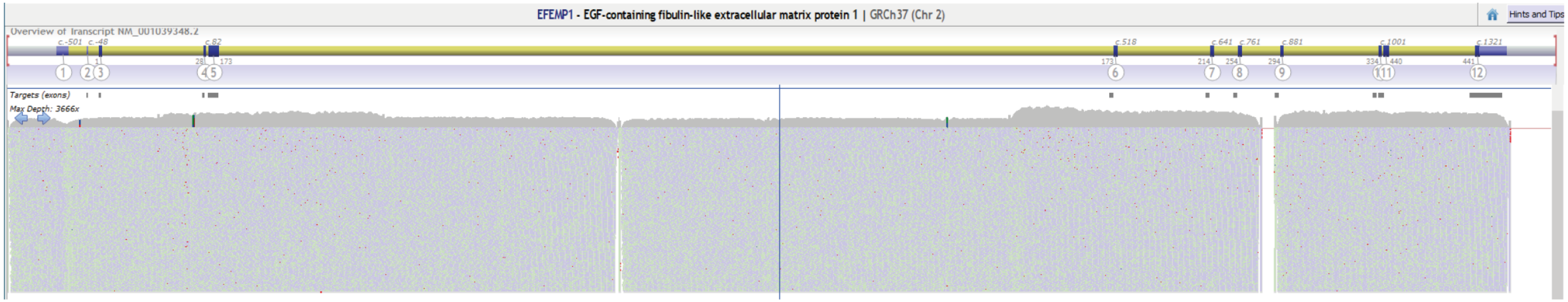
EFEMP1 - EGF-containing fibulin-like extracellular matrix protein 1 | GRCh37 (Chr 2)
Hints and Tips
Overview of Transcript NM_001039348.2
Targets (exons)
Max Depth: 3666x

E)

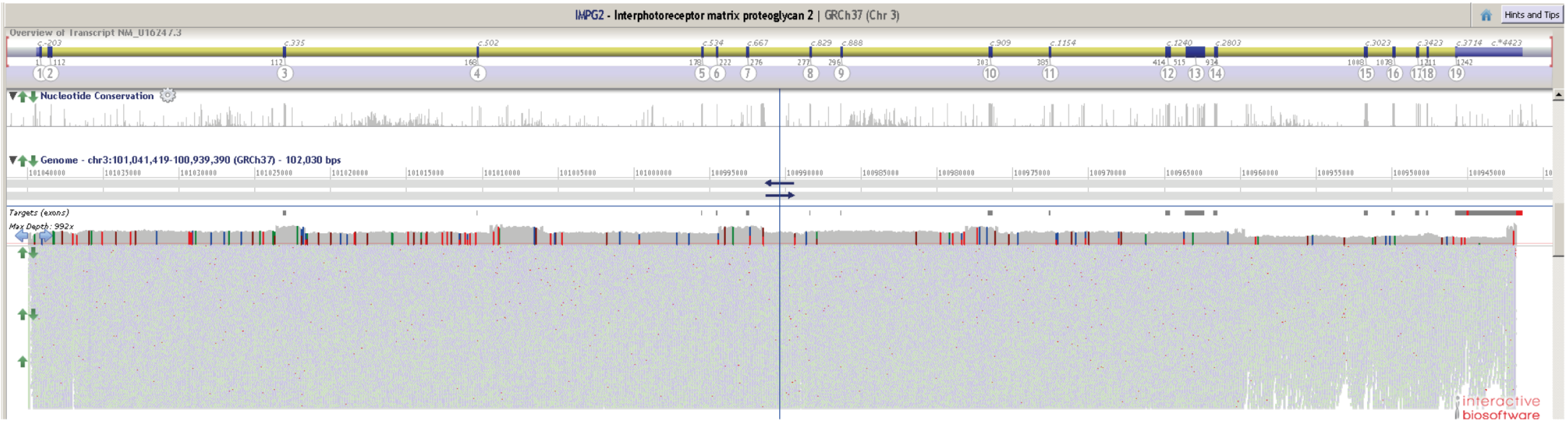
IMPG2 - Interphotoreceptor matrix proteoglycan 2 | GRCh37 (Chr 3)
Hints and Tips
Overview of Transcript NM_016247.3
Nucleotide Conservation
Genome - chr3:101,041,419-100,939,390 (GRCh37) - 102,030 bps
Targets (exons)
Max Depth: 992x
interactive biosoftware

F)

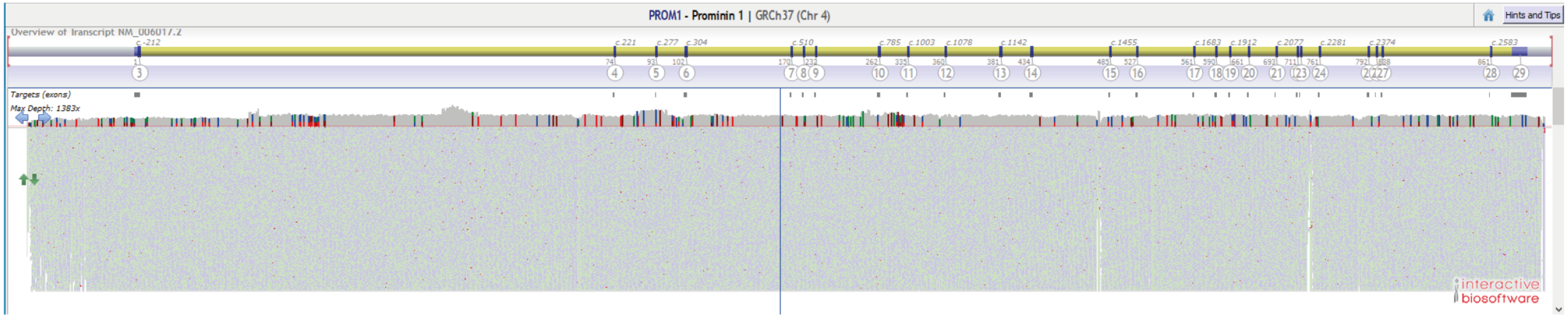
PROM1 - Prominin 1 | GRCh37 (Chr 4)
Hints and Tips
Overview of Transcript NM_006017.2
Targets (exons)
Max Depth: 1383x
interactive biosoftware

G)

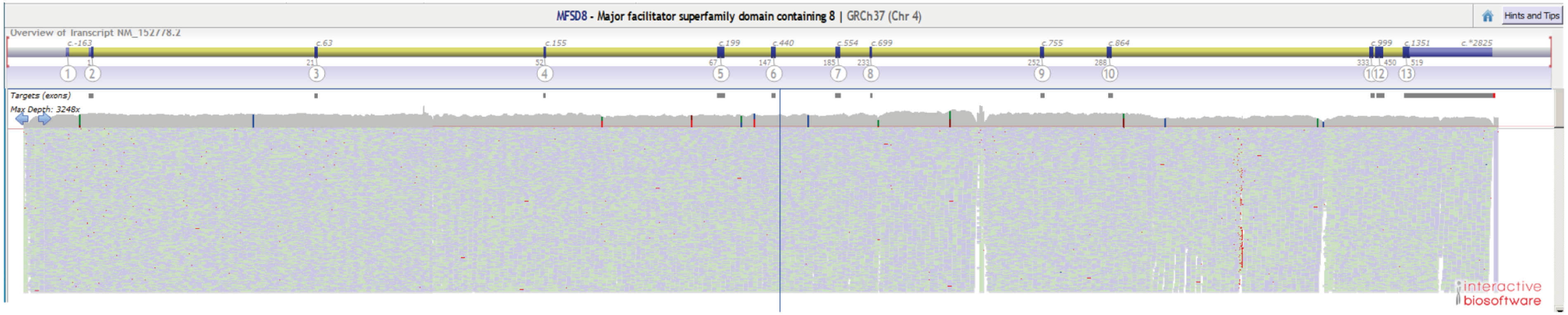
MFSD8 - Major facilitator superfamily domain containing 8 | GRCh37 (Chr 4)
Hints and Tips
Overview of Transcript NM_152778.2
Targets (exons)
Max Depth: 3248x
interactive biosoftware

H)

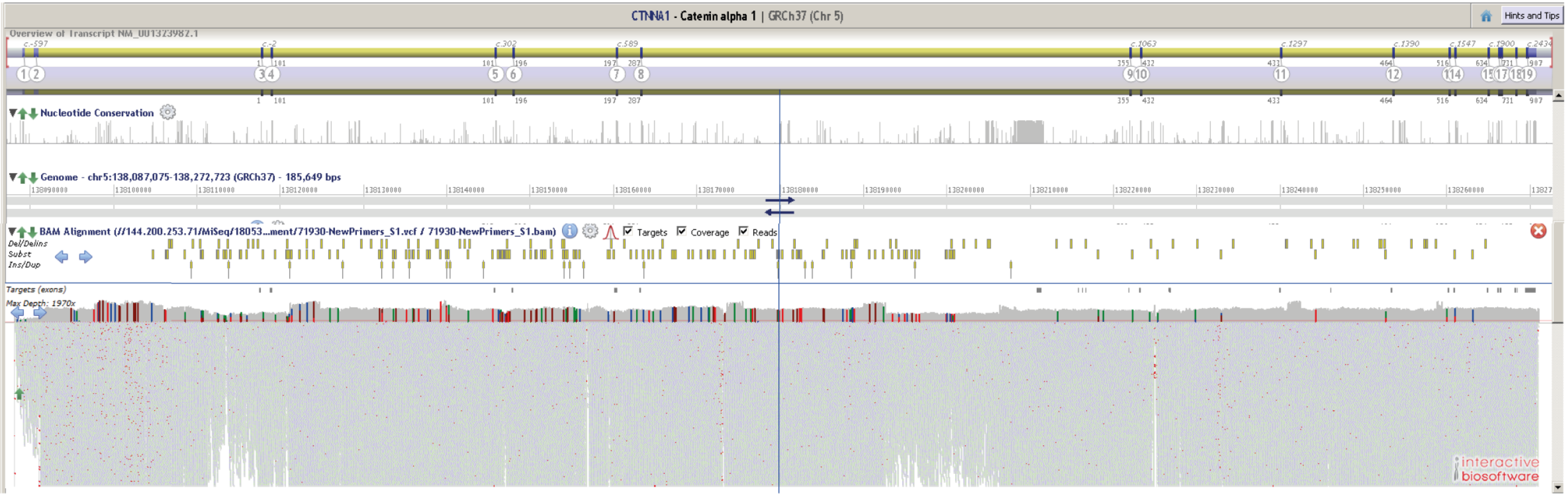

CTNNA1 - Catenin alpha 1 | GRCh37 (Chr 5)
Hints and Tips
Overview of Transcript NM_001323982.1
Nucleotide Conservation
Genome - chr5:138,087,075-138,272,723 (GRCh37) - 185,649 bps
BAM Alignment (//144.200.253.71/MiSeq/18053...ment/71930-NewPrimers_S1.vcf / 71930-NewPrimers_S1.bam)
Targets
Coverage
Reads
Del/Delins
Subst
Ins/Dup
Targets (exons)
Max Depth: 1970x
interactive biosoftware

I)

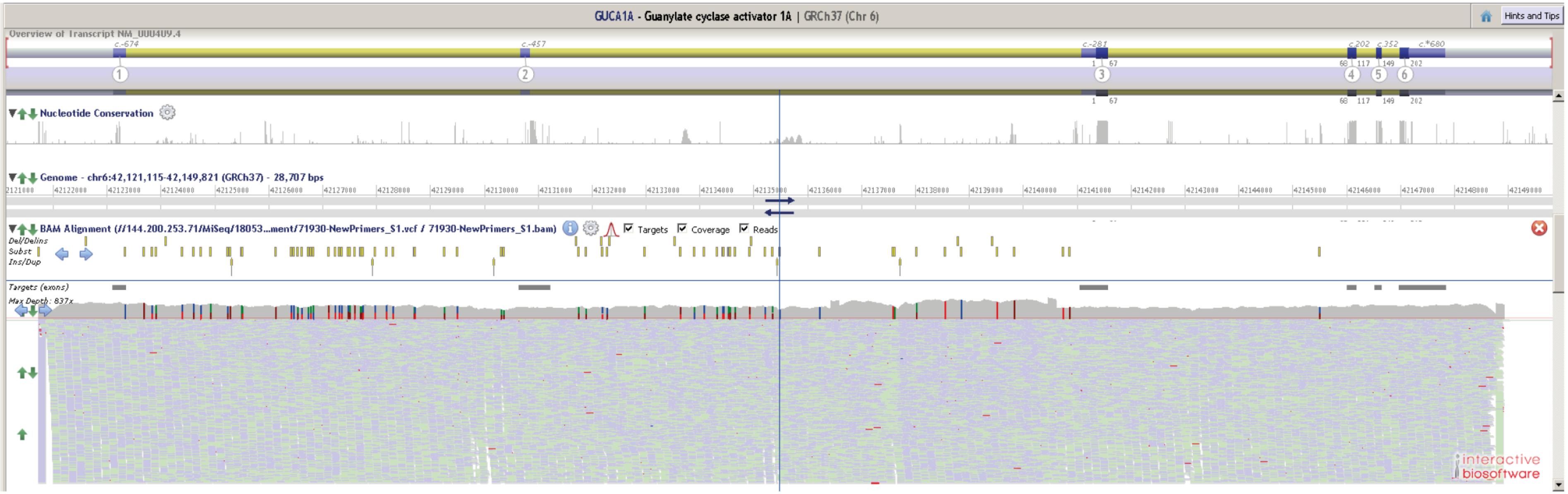
GUCA1A - Guanylate cyclase activator 1A | GRCh37 (Chr 6)
Hints and Tips
Overview of Transcript NM_000409.4
Nucleotide Conservation
Genome - chr6:42,121,115-42,149,821 (GRCh37) - 28,707 bps
BAM Alignment (//144.200.253.71/MiSeq/18053...ment/71930-NewPrimers_S1.vcf / 71930-NewPrimers_S1.bam)
Targets
Coverage
Reads
Del/Delins
Subst
Ins/Dup
Targets (exons)
Max Depth: 837x
interactive biosoftware

J)

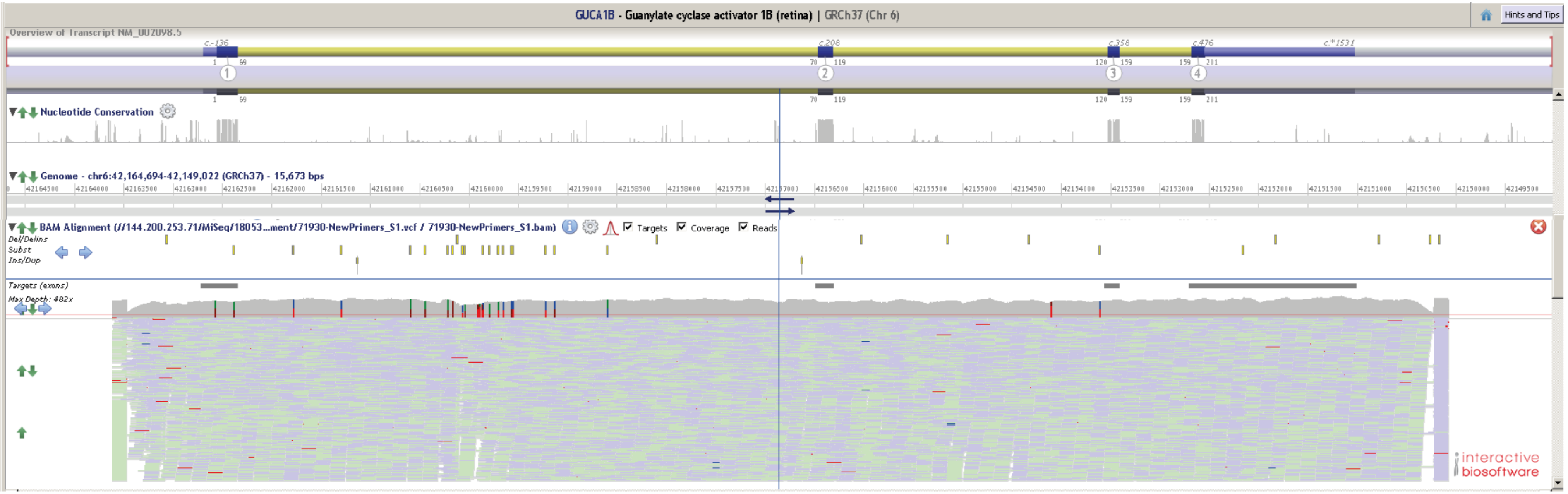
GUCA1B - Guanylate cyclase activator 1B (retina) | GRCh37 (Chr 6)
Hints and Tips
Overview of Transcript NM_002098.5
Nucleotide Conservation
Genome - chr6:42,164,694-42,149,022 (GRCh37) - 15,673 bps
Targets
Coverage
Reads
Del/Delins
Subst
Ins/Dup
Targets (exons)
Max Depth: 482x
interactive biosoftware

K)

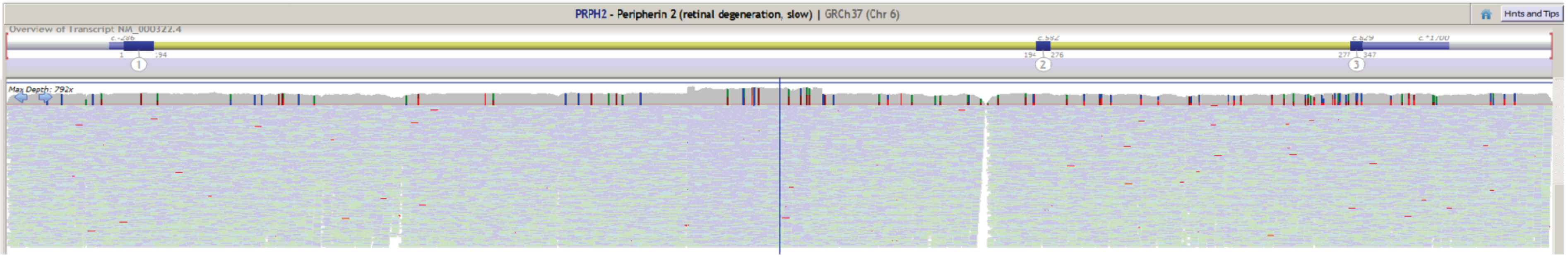
PRPH2 - Peripherin 2 (retinal degeneration, slow) | GRCh37 (Chr 6)
Hints and Tips
Overview of Transcript NM_000322.4
Max Depth: 792x

L)

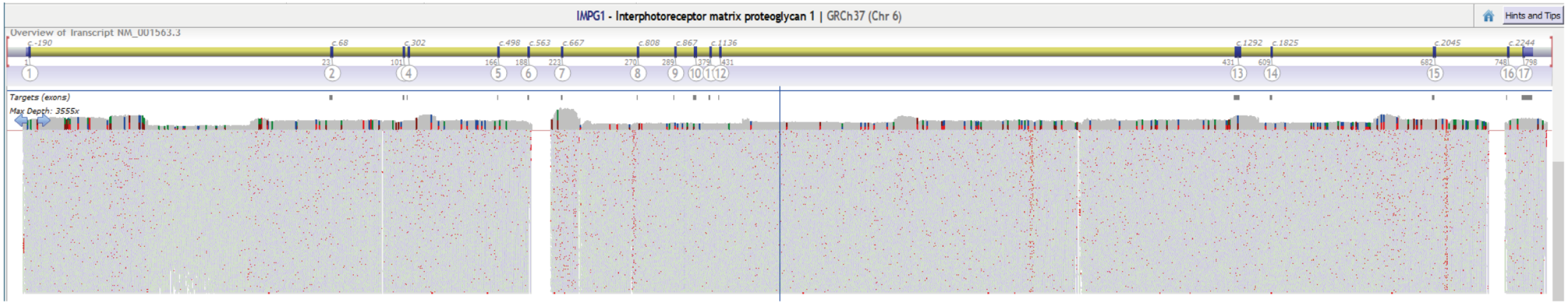
IMPG1 - Interphotoreceptor matrix proteoglycan 1 | GRCh37 (Chr 6)
Hints and Tips
Overview of Transcript NM_001563.3
Targets (exons)
Max Depth: 3555x

M)

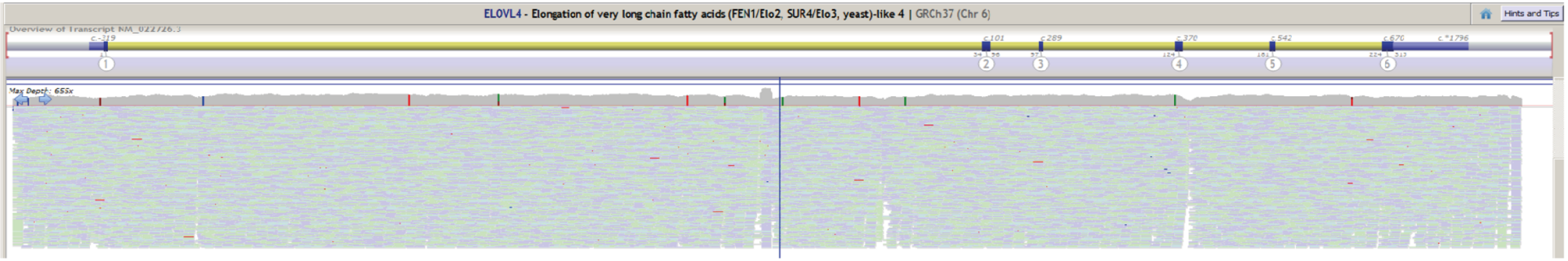
ELOVL4 - Elongation of very long chain fatty acids (FEN1/Elo2, SUR4/Elo3, yeast)-like 4 | GRCh37 (Chr 6)
Hints and Tips
Overview of Transcript NM_022726.3
Max Depth: 655x

N)

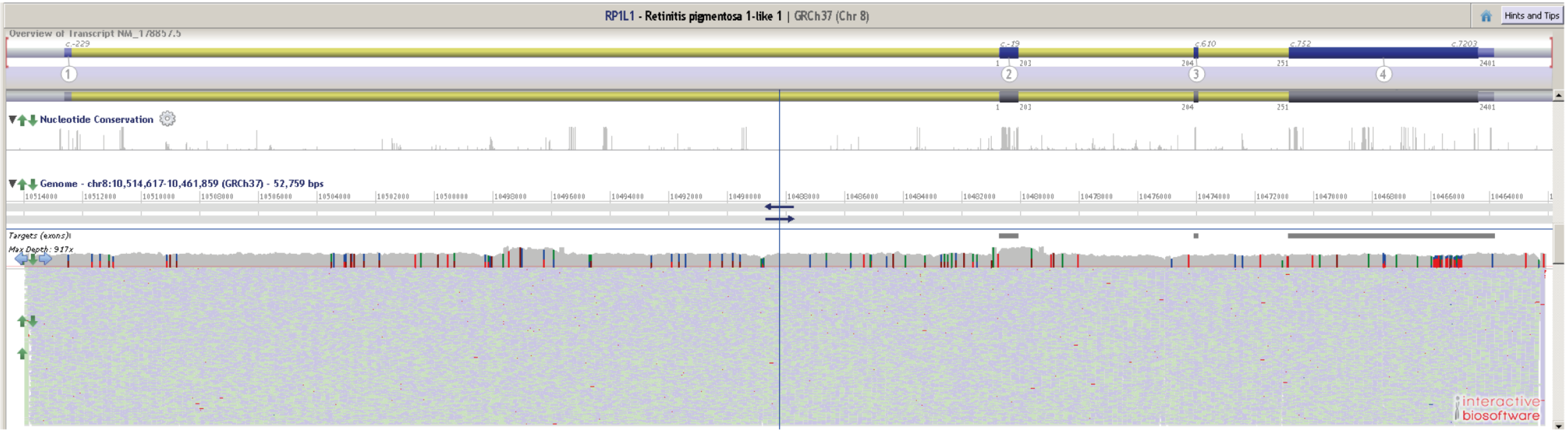
RP1L1 - Retinitis pigmentosa 1-like 1 | GRCh37 (Chr 8)
Hints and Tips
Overview of Transcript NM_178857.5
Nucleotide Conservation
Genome - chr8:10,514,617-10,461,859 (GRCh37) - 52,759 bps
Targets (exons)
Max Depth: 917x
interactive biosoftware

O)

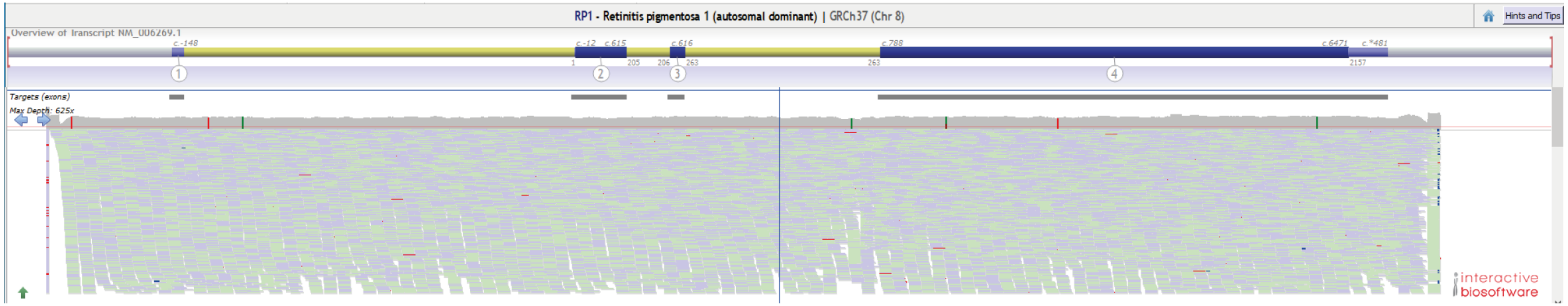
RP1 - Retinitis pigmentosa 1 (autosomal dominant) | GRCh37 (Chr 8)
Hints and Tips
Overview of Transcript NM_006269.1
Targets (exons)
Max Depth: 625x
interactive biosoftware

P)

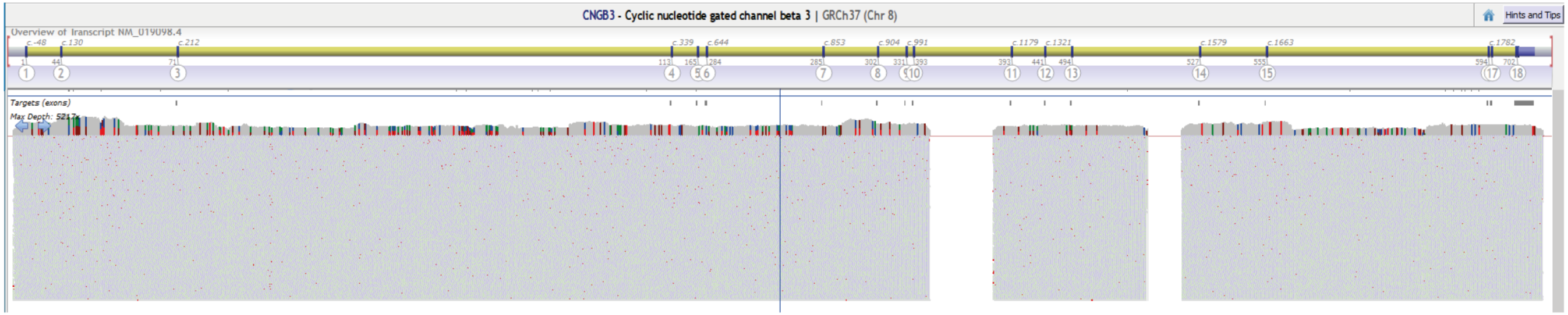
CNGB3 - Cyclic nucleotide gated channel beta 3 | GRCh37 (Chr 8)
Hints and Tips
Overview of Transcript NM_019098.4
Targets (exons)
Max Depth: 5217x

Q)

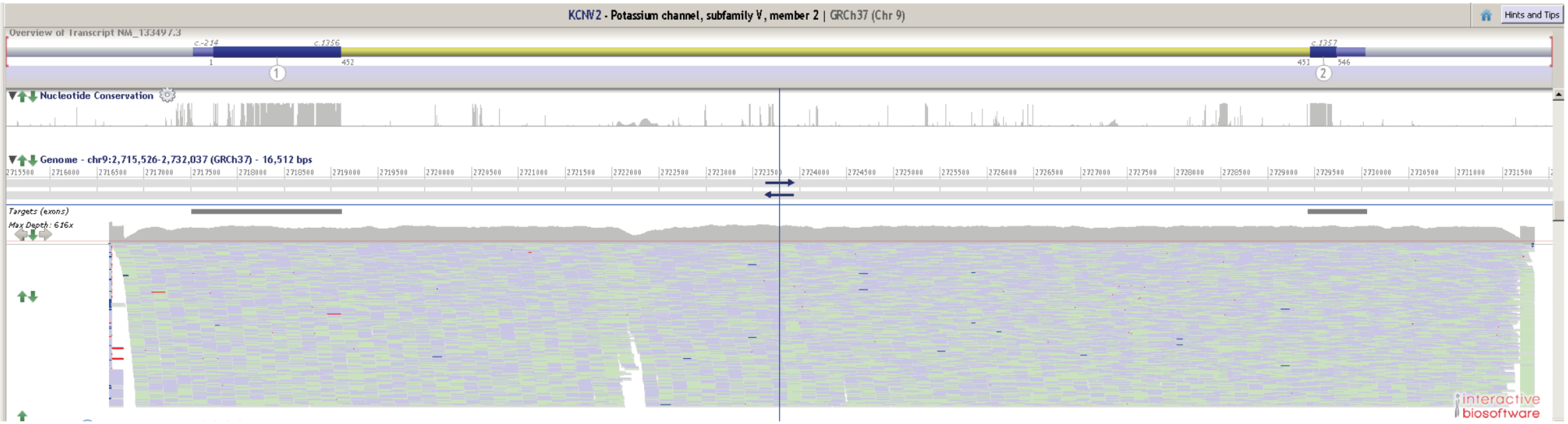
KCNV2 - Potassium channel, subfamily V, member 2 | GRCh37 (Chr 9)
Hints and Tips
Overview of Transcript NM_133497.3
Nucleotide Conservation
Genome - chr9:2,715,526-2,732,037 (GRCh37) - 16,512 bps
Targets (exons)
Max Depth: 616x
interactive biosoftware

R)

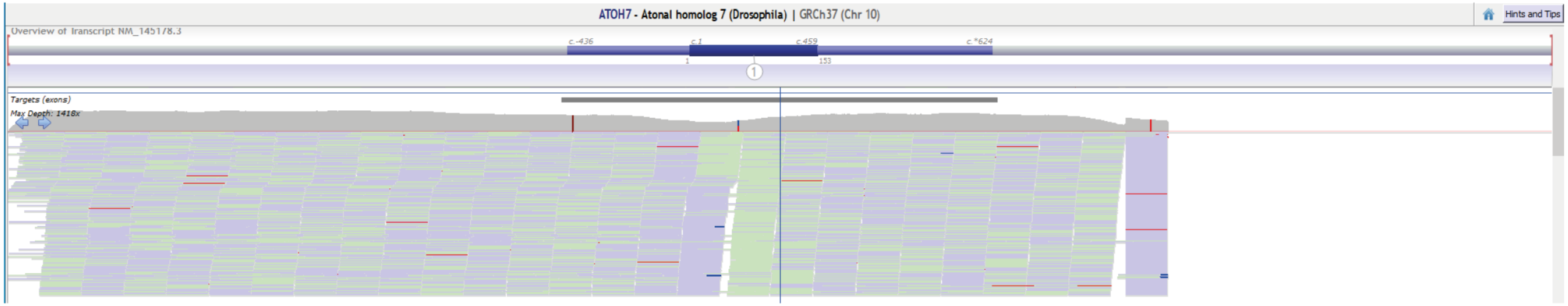
ATOH7 - Atonal homolog 7 (Drosophila) | GRCh37 (Chr 10)
Hints and Tips
Overview of Transcript NM_145178.3
c.-436
c.1
c.459
c.*624
Targets (exons)
Max Depth: 1418x

S)

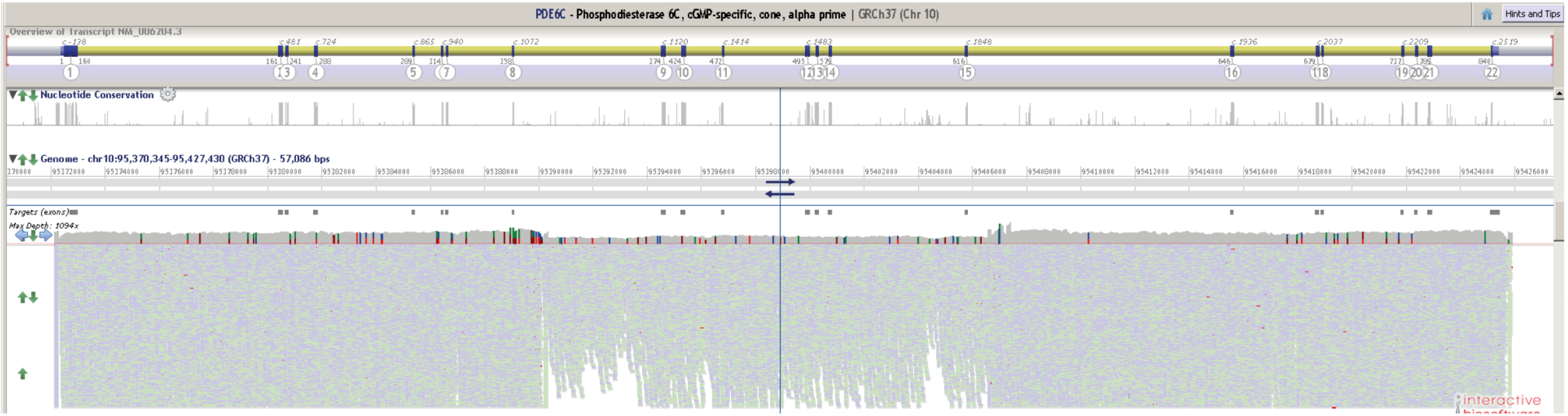
PDE6C - Phosphodiesterase 6C, cGMP-specific, cone, alpha prime | GRCh37 (Chr 10)
Hints and Tips
Overview of Transcript NM_006204.3
Nucleotide Conservation
Genome - chr10:95,370,345-95,427,430 (GRCh37) - 57,086 bps
Targets (exons)
Max Depth: 1094x
interactive

T)

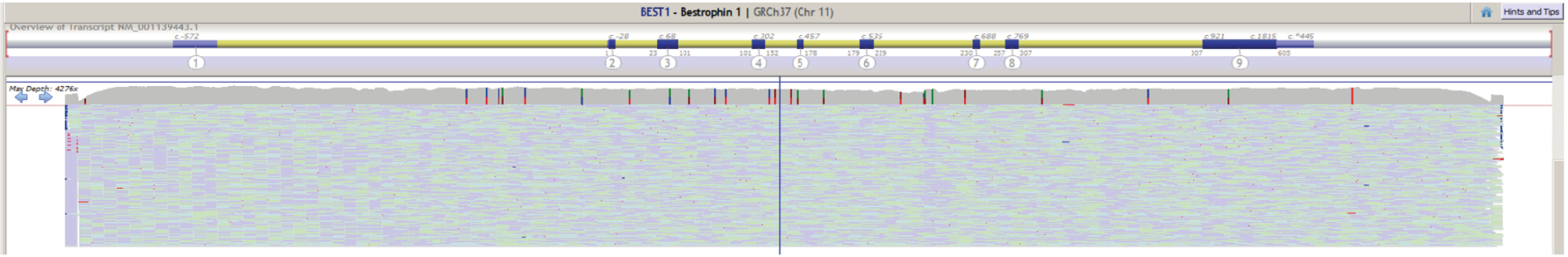
BEST1 - Bestrophin 1 | GRCh37 (Chr 11)
Hints and Tips
Overview of Transcript NM_001139443.1
Max Depth: 4276x

U)

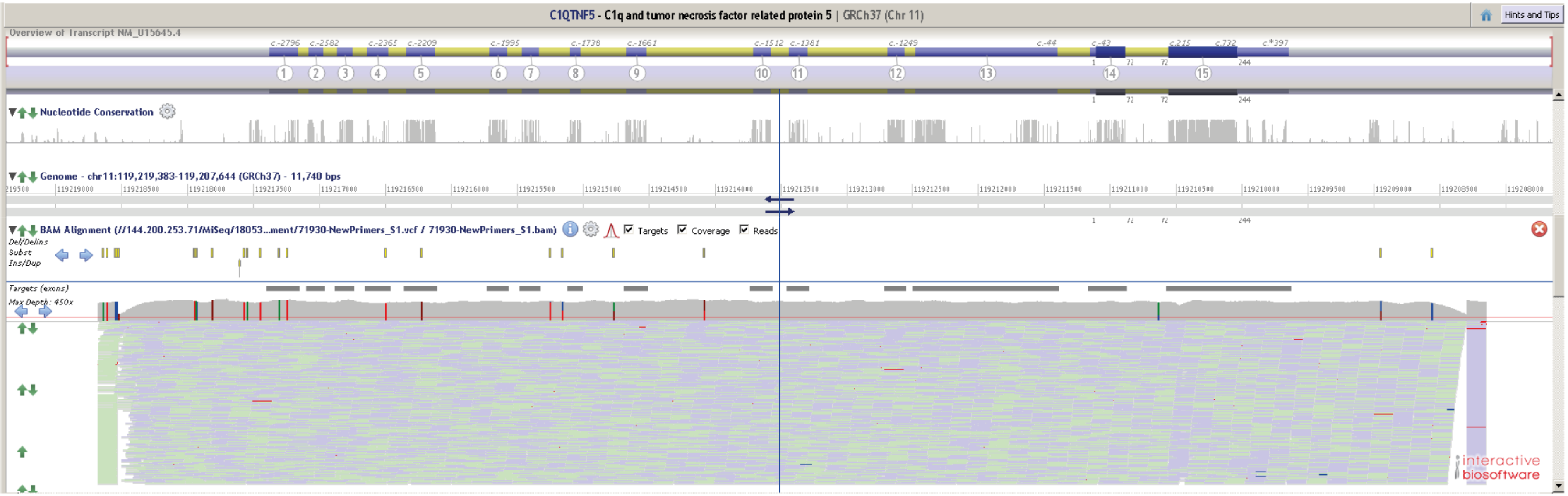
C1QTNF5 - C1q and tumor necrosis factor related protein 5 | GRCh37 (Chr 11)
Hints and Tips
Overview of Transcript NM_015645.4
Nucleotide Conservation
Genome - chr11:119,219,383-119,207,644 (GRCh37) - 11,740 bps
BAM Alignment (//144.200.253.71/MiSeq/18053...ment/71930-NewPrimers_S1.vcf / 71930-NewPrimers_S1.bam)
Targets
Coverage
Reads
Del/Delins
Subst
Ins/Dup
Targets (exons)
Max Depth: 450x
interactive biosoftware

V)

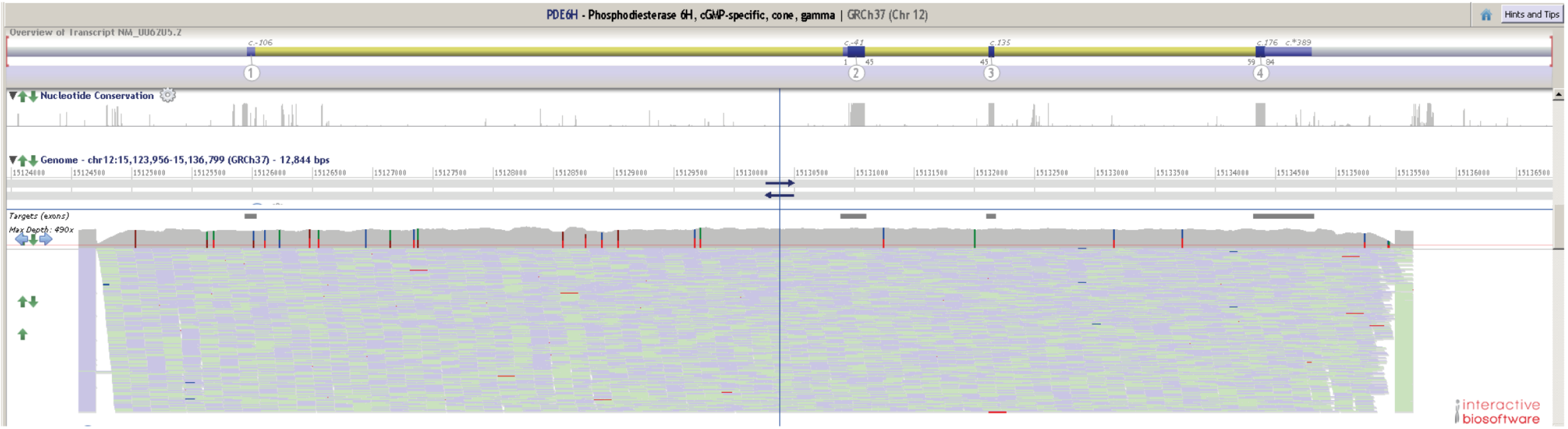
PDE6H - Phosphodiesterase 6H, cGMP-specific, cone, gamma | GRCh37 (Chr 12)
Hints and Tips
Overview of Transcript NM_006205.2
Nucleotide Conservation
Genome - chr12:15,123,956-15,136,799 (GRCh37) - 12,844 bps
Targets (exons)
Max Depth: 490x
interactive
biosoftware

W)

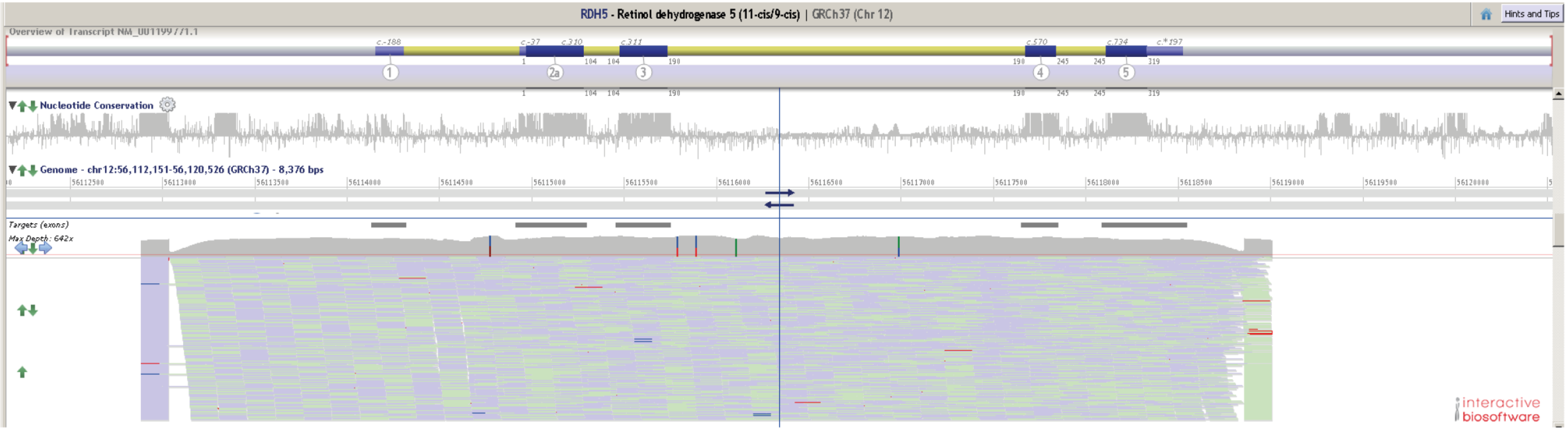
RDH5 - Retinol dehydrogenase 5 (11-cis/9-cis) | GRCh37 (Chr 12)
Hints and Tips
Overview of Transcript NM_001199771.1
Nucleotide Conservation
Genome - chr12:56,112,151-56,120,526 (GRCh37) - 8,376 bps
Targets (exons)
Max Depth: 642x
interactive
biosoftware

X)

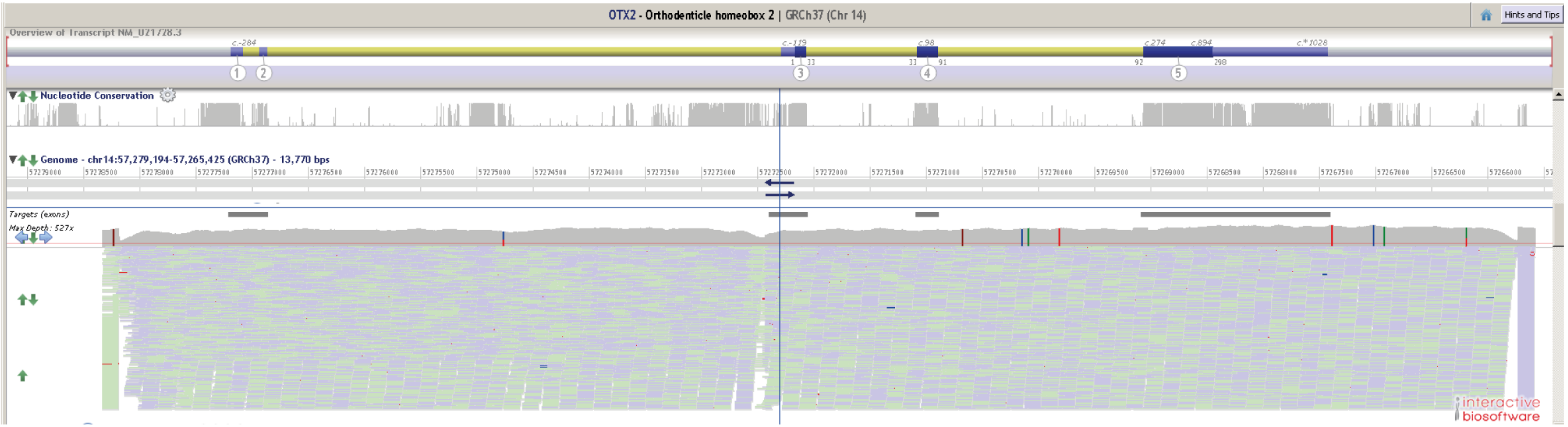
OTX2 - Orthodenticle homeobox 2 | GRCh37 (Chr 14)
Hints and Tips
Overview of Transcript NM_021728.3
Nucleotide Conservation
Genome - chr14:57,279,194-57,265,425 (GRCh37) - 13,770 bps
Targets (exons)
Max Depth: 527x
interactive biosoftware

Y)

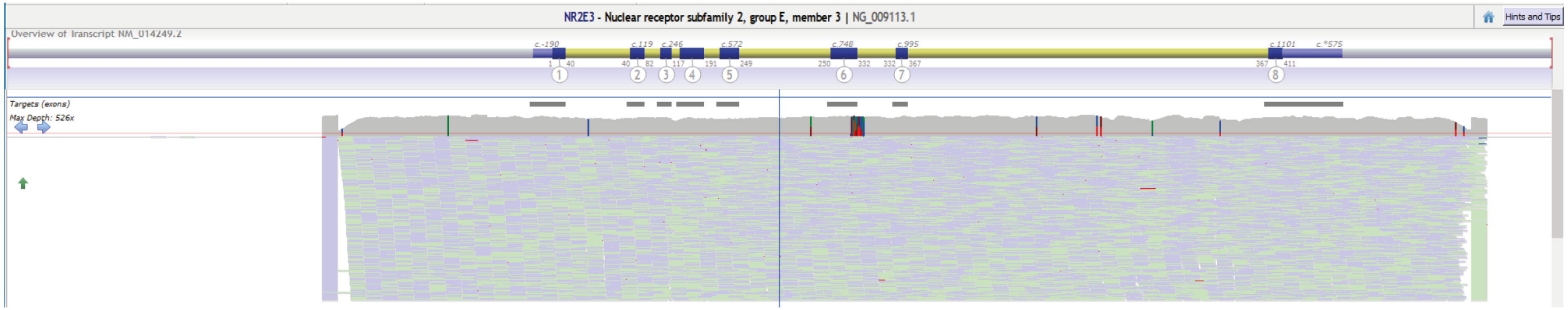
NR2E3 - Nuclear receptor subfamily 2, group E, member 3 | NG_009113.1
Hints and Tips
Overview of Transcript NM_014249.2
Targets (exons)
Max Depth: 526x

Z)

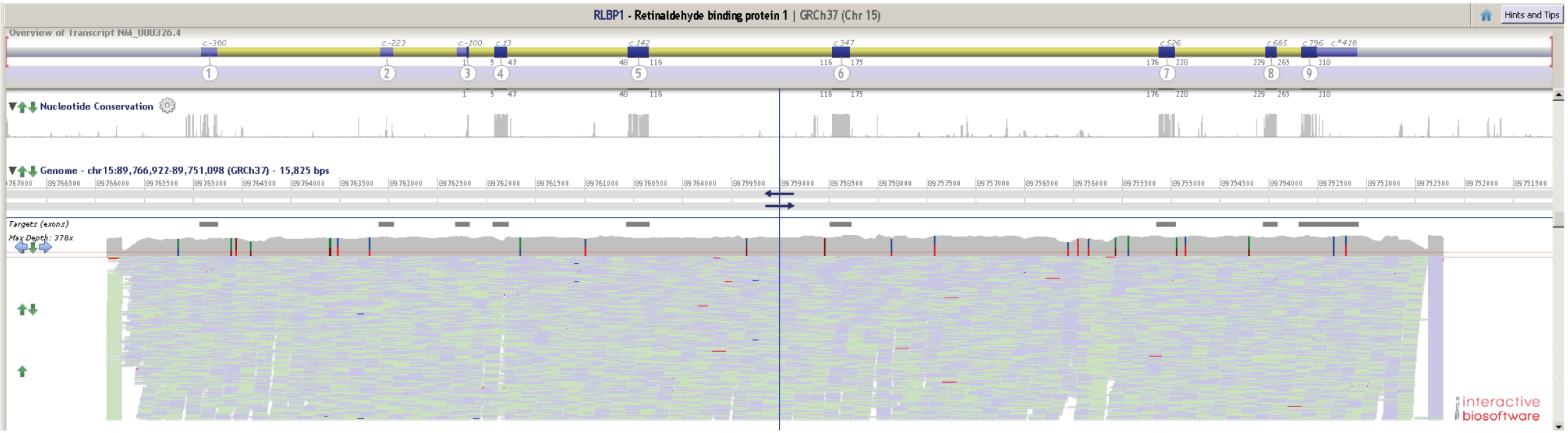

RLBP1 - Retinaldehyde binding protein 1 | GRCh37 (Chr 15)
Hints and Tips
Overview of Transcript NM_000326.4
Nucleotide Conservation
Genome - chr15:89,766,922-89,751,098 (GRCh37) - 15,825 bps
Targets (exons)
Max Depth: 378x
interactive biosoftware


AA)

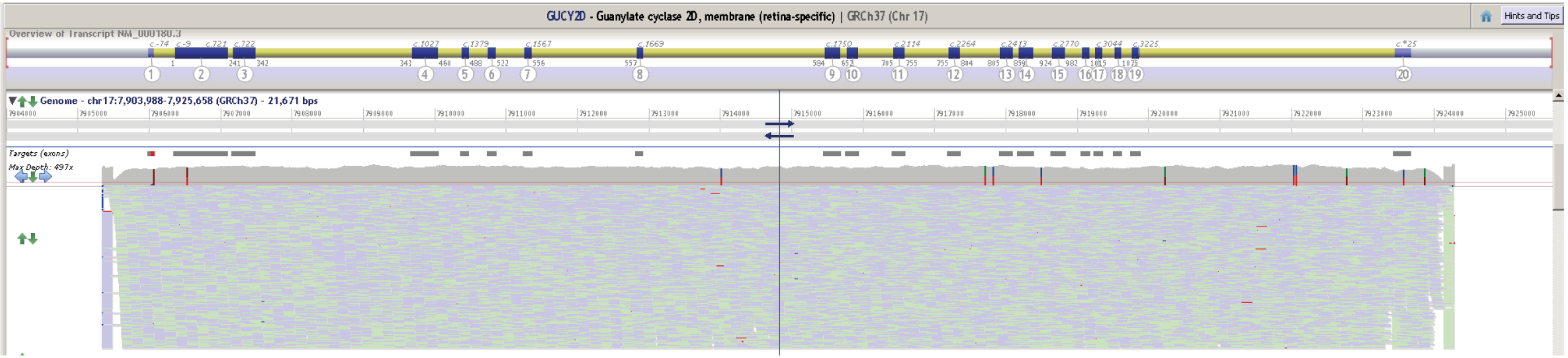

GUCY2D - Guanylate cyclase 2D, membrane (retina-specific) | GRCh37 (Chr 17)
Hints and Tips
Overview of Transcript NM_000180.3
Genome - chr17:7,903,988-7,925,658 (GRCh37) - 21,671 bps
Targets (exons)
Max Depth: 497x

BB)

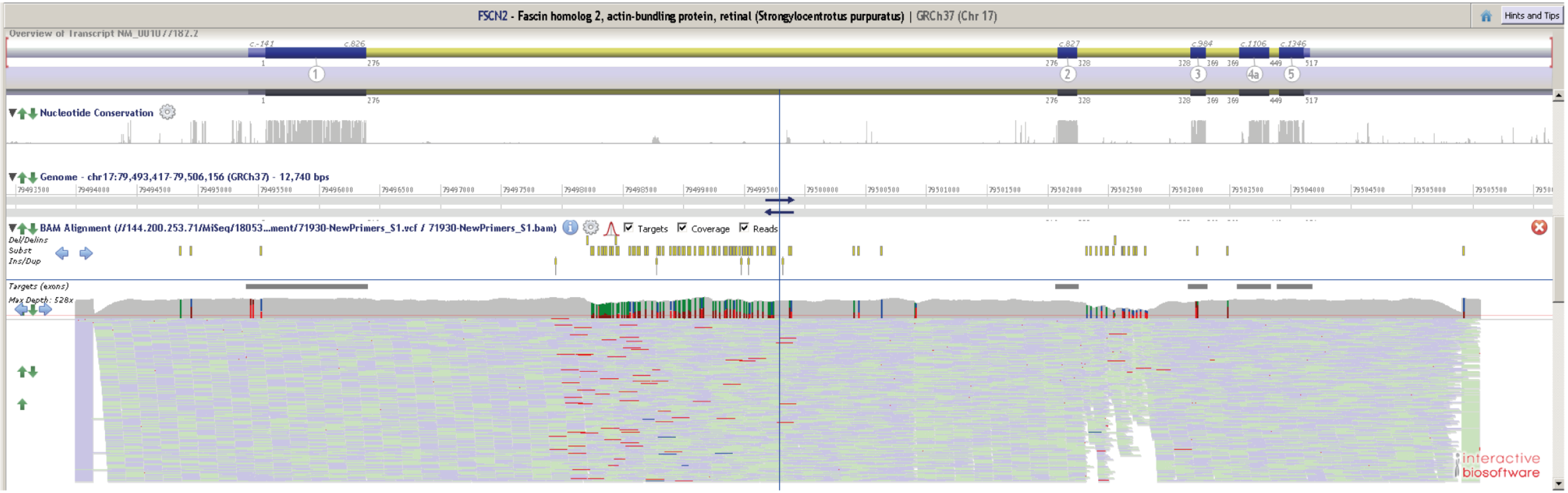

FSCN2 - Fascin homolog 2, actin-bundling protein, retinal (Strongylocentrotus purpuratus) | GRCh37 (Chr 17)
Hints and Tips
Overview of Transcript NM_001077182.2
Nucleotide Conservation
Genome - chr17:79,493,417-79,506,156 (GRCh37) - 12,740 bps
BAM Alignment (//144.200.253.71/MiSeq/18053...ment/71930-NewPrimers_S1.vcf / 71930-NewPrimers_S1.bam)
Targets
Coverage
Reads
Del/Delins
Subst
Ins/Dup
Targets (exons)
Max Depth: 528x
interactive biosoftware

CC)

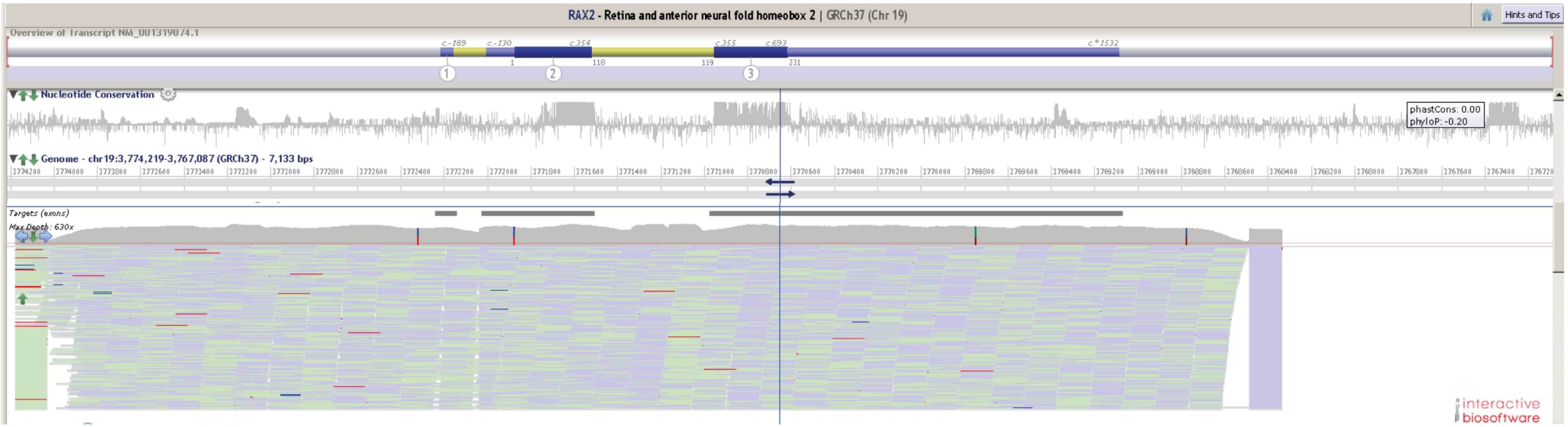

RAX2 - Retina and anterior neural fold homeobox 2 | GRCh37 (Chr 19)
Hints and Tips
Nucleotide Conservation
phastCons: 0.00
phyloP: -0.20
Genome - chr19:3,774,219-3,767,087 (GRCh37) - 7,133 bps
Targets (exons)
Max Depth: 630x
interactive biosoftware


DD)

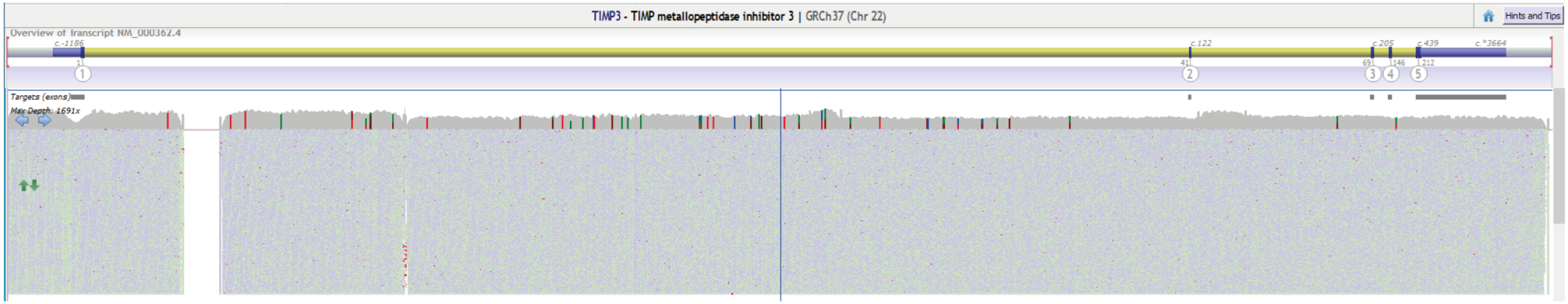

TIMP3 - TIMP metallopeptidase inhibitor 3 | GRCh37 (Chr 22)
Hints and Tips
Targets (exons)
Max Depth: 1691x

EE)

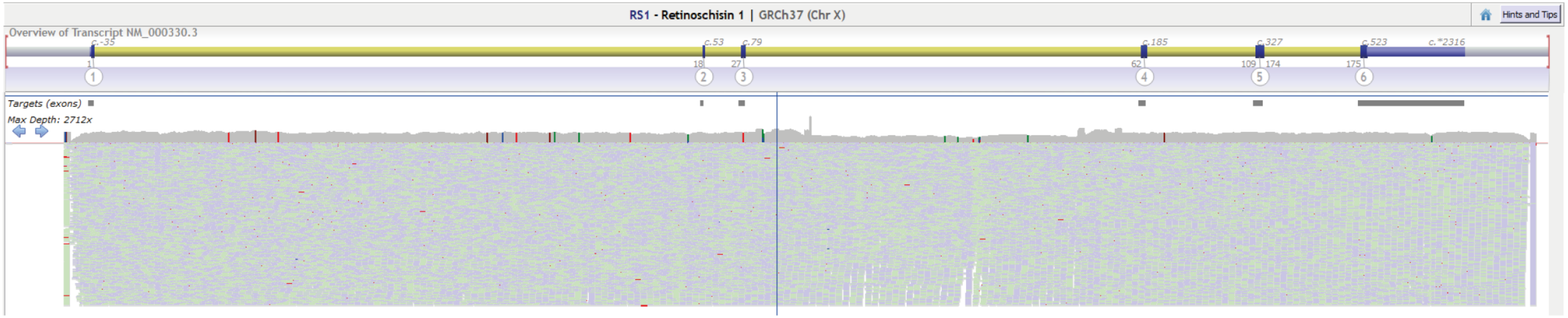
RS1 - Retinoschisin 1 | GRCh37 (Chr X)
Hints and Tips
Overview of Transcript NM_000330.3
Targets (exons)
Max Depth: 2712x

FF)

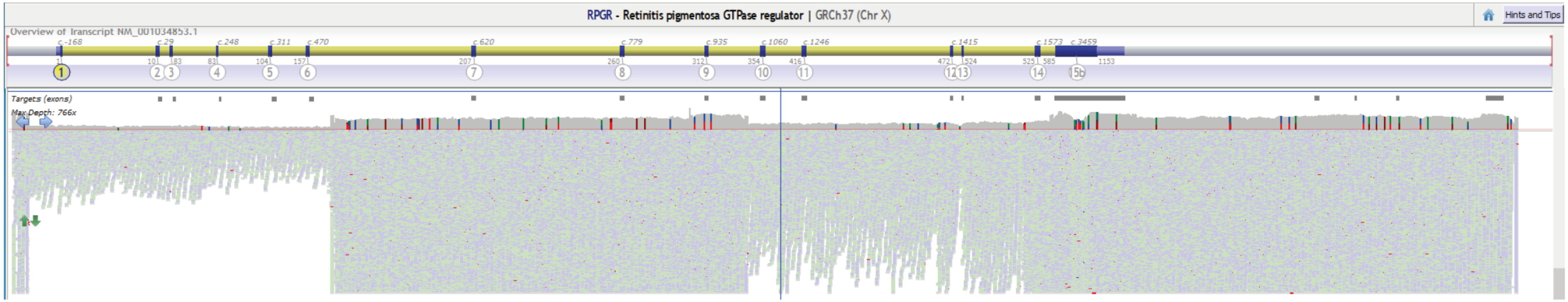
RPGR - Retinitis pigmentosa GTPase regulator | GRCh37 (Chr X)
Hints and Tips
Overview of Transcript NM_001034853.1
Targets (exons)
Max Depth: 766x

GG)

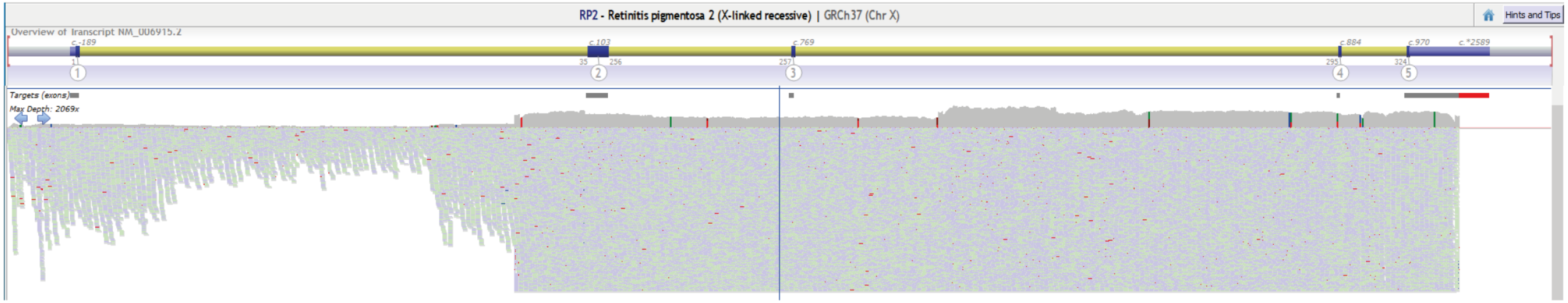

RP2 - Retinitis pigmentosa 2 (X-linked recessive) | GRCh37 (Chr X)
Hints and Tips
Overview of Transcript NM_006915.2
c.-189
c.103
c.769
c.884
c.970
c.*2589
1
35
256
257
295
324
Targets (exons)
Max Depth: 2069x

HH)

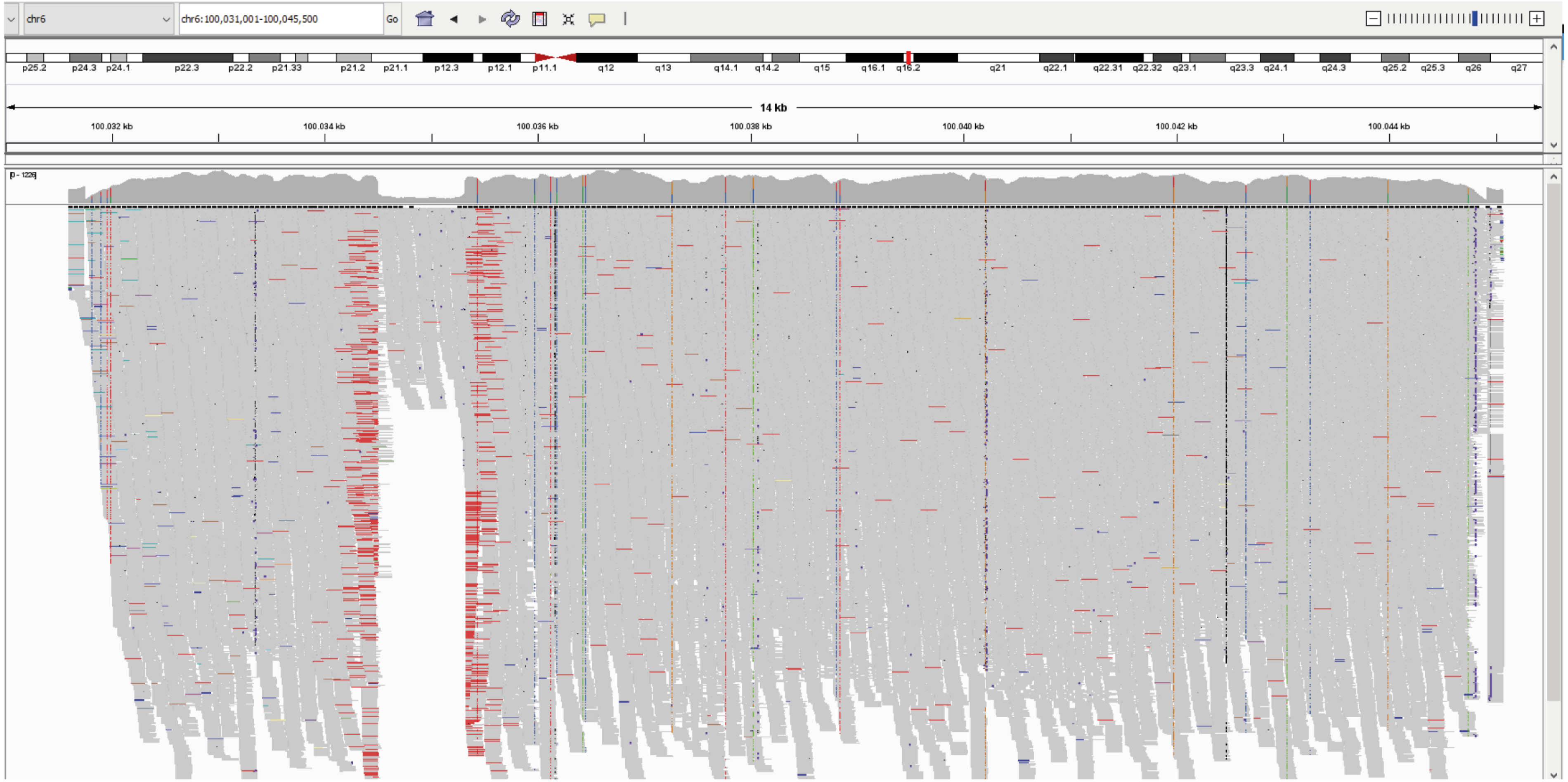
chr6
chr6:100,031,001-100,045,500
Go
p25.2 p24.3 p24.1 p22.3 p22.2 p21.33 p21.2 p21.1 p12.3 p12.1 p11.1 q12 q13 q14.1 q14.2 q15 q16.1 q16.2 q21 q22.1 q22.31 q22.32 q23.1 q23.3 q24.1 q24.3 q25.2 q25.3 q26 q27
14 kb
100.032 kb
100.034 kb
100.036 kb
100.038 kb
100.040 kb
100.042 kb
100.044 kb
