## Supplementary Figure S2-4 for "Long-range PCR-based NGS applications to diagnose Mendelian retinal diseases" (from another version of this preprint: version 1, 2020-12-02)

**Figure S2-4.** *KCNV2*, *RS1*, and *ABCA4* deletions characterization and chromosomal context.

**Figure S2**. *ABCA4* NC_000001.10:g.(94507656_94507700)_(94511700_94511710)del (3999-4033 bp)
A) Chromosomal overview and zoom into the *ABCA4* locus. The red bar highlights the location and size of the deletion.
B) Magnification of the region encompassing exons 19 to 23 of *ABCA4*. The red bar shows the relative position and size of the deletion. The black bars mark the location of repeat elements from RepeatMasker. Two Alu elements are present in close proximity with both the proximal and distal breakpoints.
C) NGS alignment of the proximal breakpoint region (left, red square in Figure S1B) and of the distal breakpoint region (right, red square in Figure S1B). The alignment shows the location of variants with different colors. The relative location of the Alu elements on the proximal and distal side is highlighted with a blue and an orange line, respectively. The green line shows a 10 bp-long homologous sequence present on both sides of the deleted region.

**Figure S3**. *KCNV2* NC_000009.11:g.2716981_2787016del (70036 bp)
A) Chromosomal overview and zoom into the *KCNV2* and neighbouring genes region. The figure highlights the deletions published previously by Wissinger et al. in 2011 (blue bars), as well as the position of that identified in the present study (red bar).
B) CytoSNP-850K BeadChip results for the *KCNV2* region (corresponding to the red square in Figure S2A). The B-allele frequency plot is shown on top and the corresponding log R ratio plot is displayed below.
C) NGS alignment results for the breakpoint regions. On the left, the 5' side breakpoint alignments are shown. The sequence is illustrated underneath the reads; the blue line underneath the sequence highlights the part of the junction region that originates from the proximal side of the deletion. The green line marks the 3 bp-long homologous sequence that is present on both sides of the deleted region. Finally, the orange line shows the part of the sequence coming from the distal side.
D) Electropherogram of the junction fragment for the index patient. The resulting sequence is reported above the peaks. The fragments of the proximal, distal and the shared homologous sequences are highlighted with blue, orange, and green lines, respectively.

**Figure S4**. *RS1* NC_000023.10:g.18675498_18676502del (1005 bp)
A) Chromosomal overview and zoom into the *RS1* and neighbouring gene region. The red bar shows the position and size of the deletion.
B) NGS alignment to the red square region in Figure S3A.
C) Magnification of the 5' (left) and 3' (right) breakpoint regions. The corresponding sequence is reported underneath the alignment figure. The blue line under the sequence shows the part of the junction region that originates from the proximal side of the deletion. The green line highlights the 4 bp-long homologous sequence that is present on both sides of the deleted region. The orange line marks the part of the sequence coming from the distal side.

### *ABCA4*

NC_000001.10:g.(94507656_94507700)_(94511700_94511710)del (3999-4033 bp)

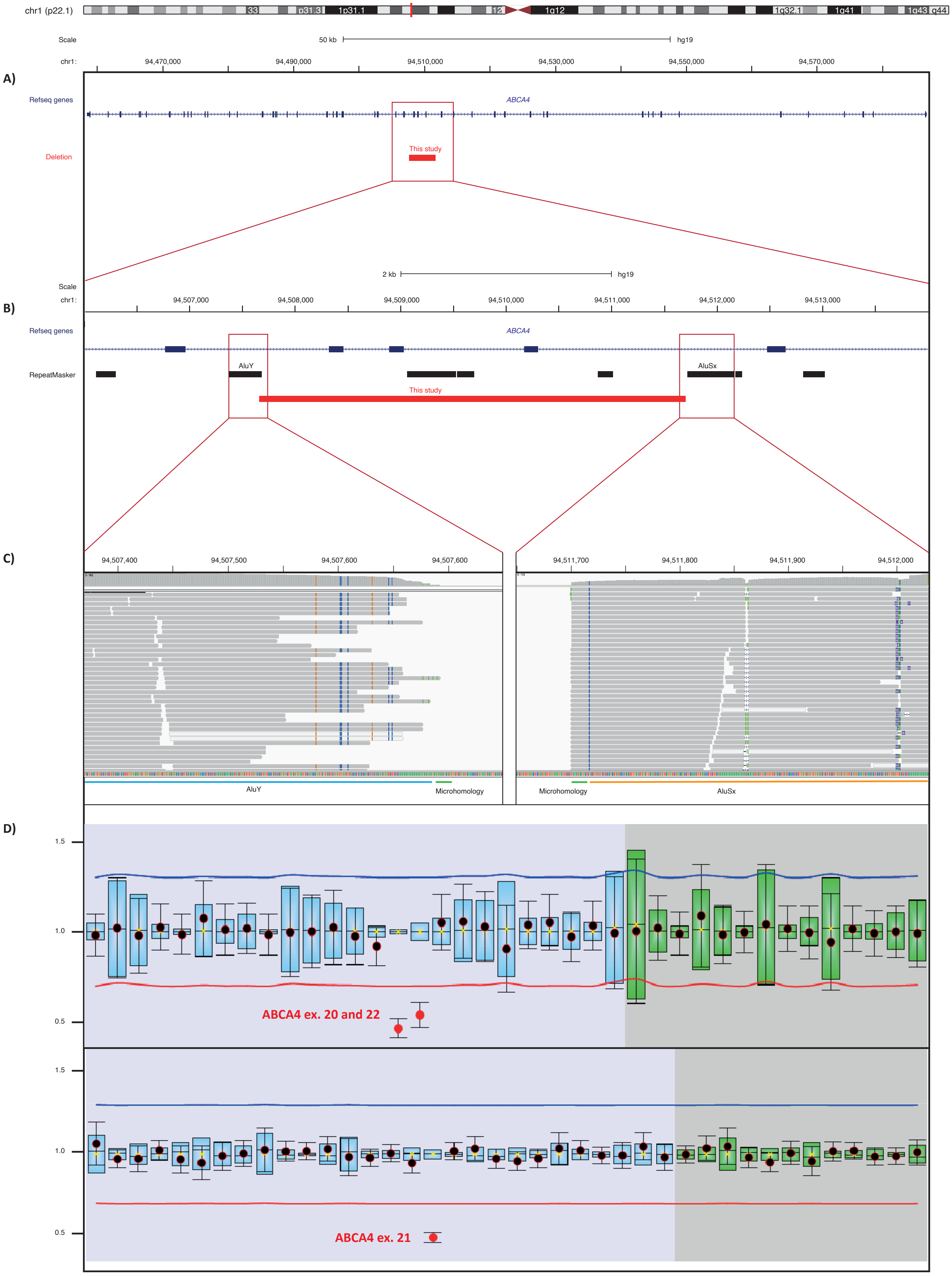

***KCNV2***

NC_000009.11:g.2716981_2787016del (70'036 bp)

NM_133497.3:c.-759_*57289del

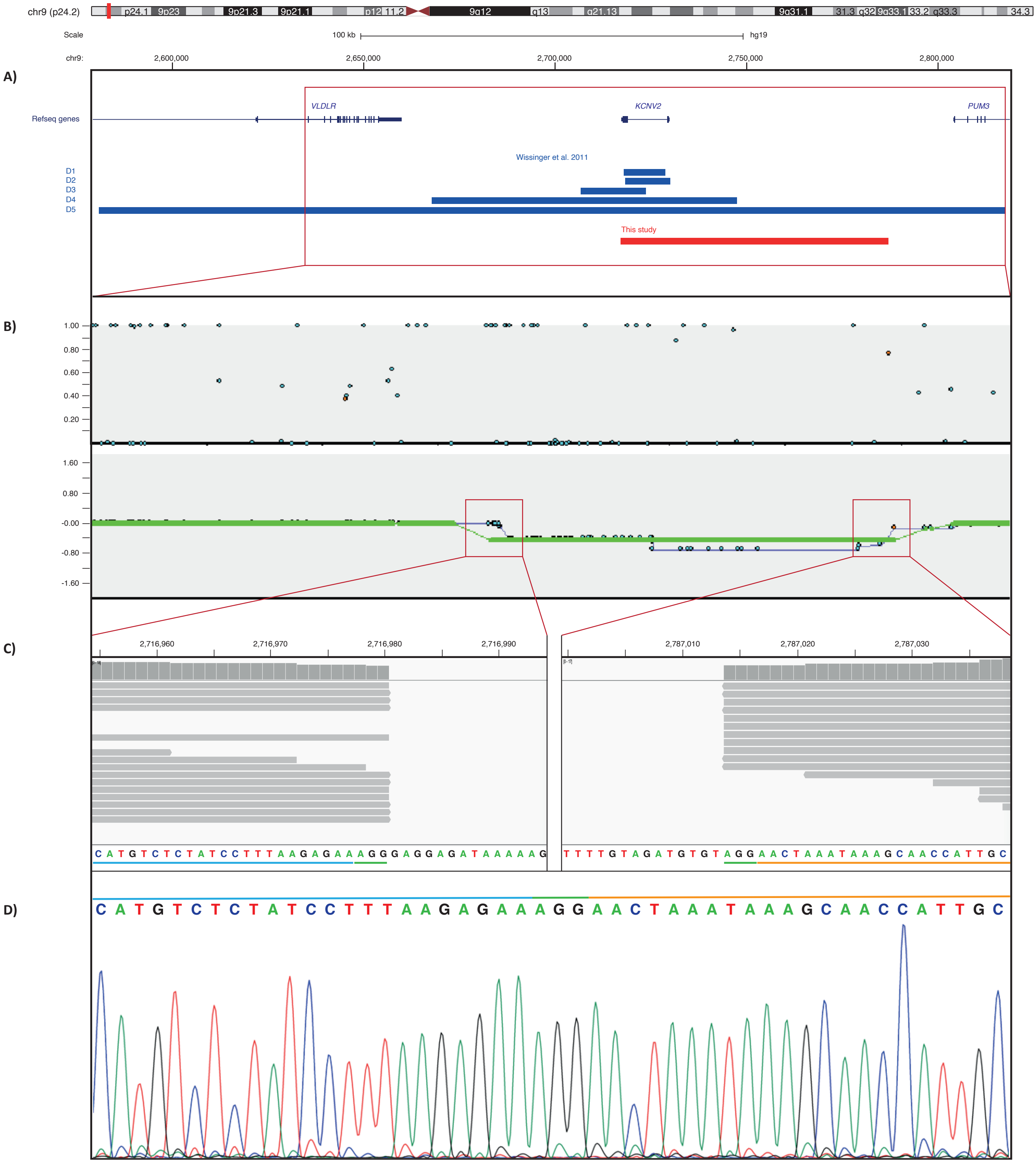

## *RS1*

NC_000023.10:g.18675498_18676502del (1005 bp)

NM_000330.3:c.53-717_78+262del

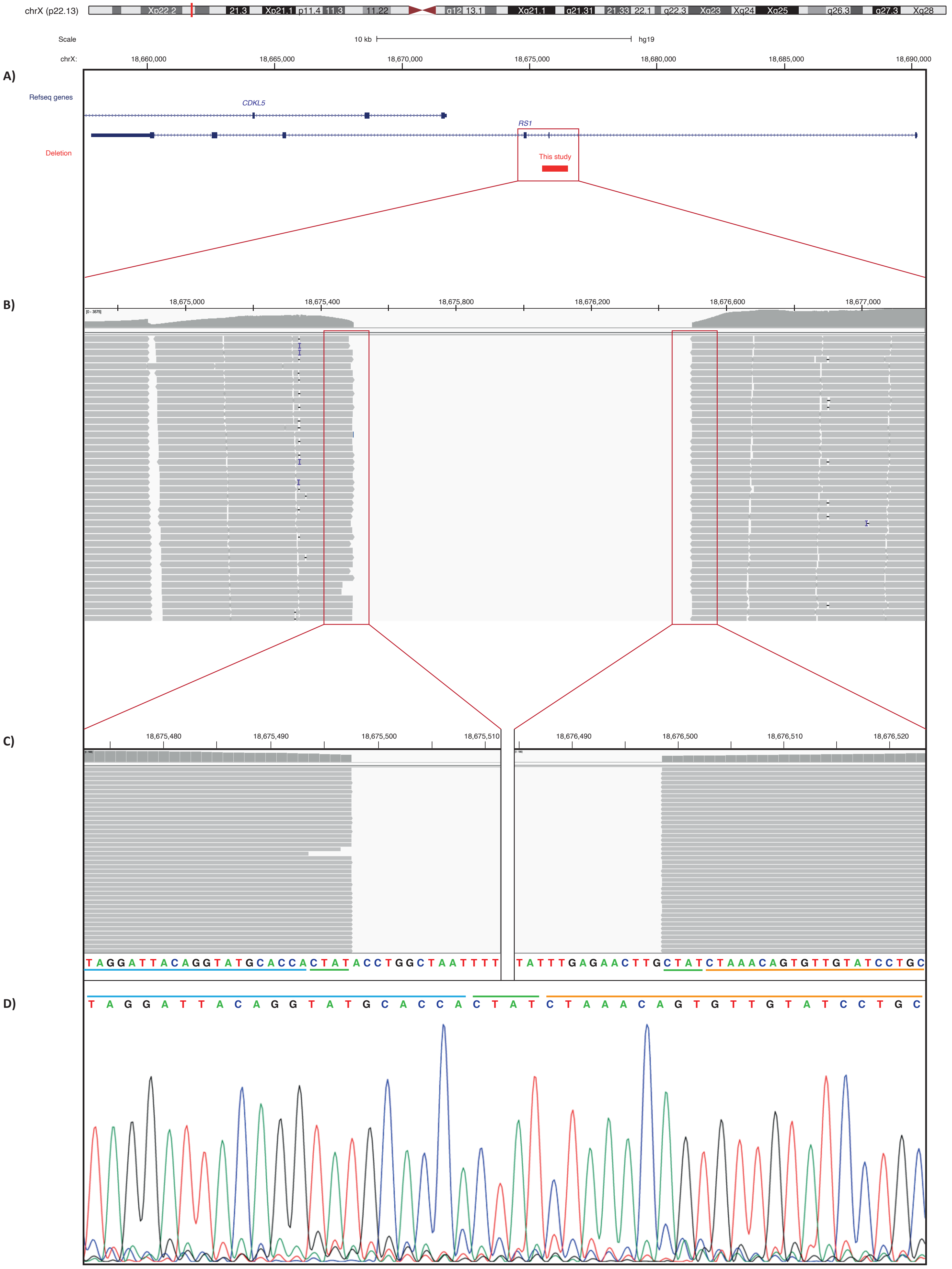
