## Supplementary Text S1 for "Long-range PCR-based NGS applications to diagnose Mendelian retinal diseases" (from another version of this preprint: version 1, 2020-12-02)

**Long-range PCR library prep protocol**

1. **gDNA dilution**
   a. Normalize all gDNA templates to the same concentration (10 ng/µl) in IDTE buffer (TE buffer pH 8.0) or ddH_2_O
   b. Check DNA concentration with NanoDrop
2. **Long-range PCR**
   a. Complete the following table listing all PCR reactions to be done with the corresponding templates

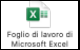

b. Insert the total number of reactions in the following table (calculate a 10% excess for pipetting errors):

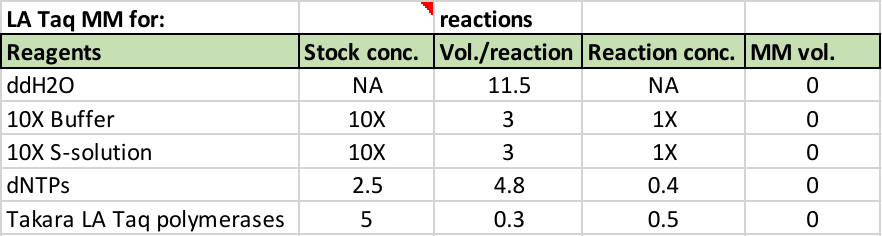

c. Thaw 10X Buffer, 10X S-solution, dNTPs, region-specific primers, and the selected gDNA templates at RT
d. Prepare the universal MM without primers according to the table above
e. Mix well by pipetting up and down
f. Prepare region-specific master mixes from the universal MM, if necessary:
   i. Complete the primer-specific MM table according to the number of reactions needed (calculate a 10% excess for pipetting errors) for each MM needed

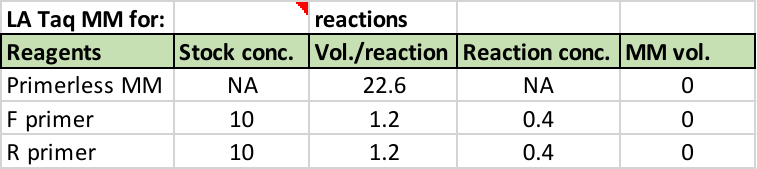

g. From the primerless MM, create primer-specific Master Mixes according to the mixes above
h. Mix well each primer-specific MM by pipetting up and down
i. For small numbers of reactions, prepare an appropriate amount of 0.2 ml PCR tubes and label them
j. Otherwise, plan how to distribute the reactions in a PCR plate:

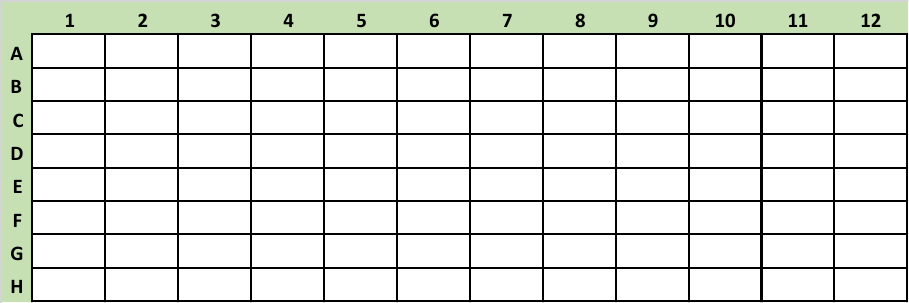

k. Distribute 25 µl of primer-specific MM to the corresponding tubes or wells
l. Add 5 µl of the template gDNA dilutions (50 ng of DNA in total)
m. Mix by pipetting up and down
n. Place the PCR tubes/plate on the thermal cycler with the **Long-Range 2-Step PCR** program:

| Stage | Temperature (Lid: 105°C) | Time | Number of cycles |
| --- | --- | --- | --- |
| Initial denaturation | 94°C | 2 min | 1 |
| Denaturation, annealing and elongation | 98°C | 10 s | 35 |
|  | 68°C | 12 min |  |
| Final elongation | 72°C | 10 min | 1 |
| Hold | 4°C | ∞ |  |

3. **Electrophoresis**
   a. Preparation:
      i. Depending on how many PCR reactions there are, decide on a particular gel size with corresponding comb
      ii. Prepare the chosen gel chamber by sealing the sides with tape so that the liquid gel solution is confined to the chamber
   b. Prepare a 0.6% agarose gel for the chosen quantity
      i. E.g. for a 100-ml gel pour the following mix into a 250-ml Erlenmeyer flask:

| Reagent | Quantity |
| --- | --- |
| Agarose powder | 0.6 g |
| 1X TAE buffer | 100 ml |

c. Heat the mix in a microwave oven until it starts boiling and the solution is completely clear
d. Wait for the mixture to cool down and/or speed up the cooling process by swirling the solution in the Erlenmeyer flask under cold running tap water until it reaches a temperature of approximately 60-70˚C
e. Transfer the solution into an EthBr-dedicated Erlenmeyer
f. Add an appropriate amount of EthBr reagent
   i. E.g. for a 100-ml gel add 5 µl
g. Mix gently by swirling the solution
h. Carefully pour the gel solution onto the previously taped gel chamber
i. Ensure no bubbles remain
j. Add the appropriate comb(s)
k. Let the gel cool down and solidify for approximately 30 minutes at RT
l. Put the gel chamber in the cold room (or in the fridge) for approximately one hour
   i. Before taking the gel out of the cold room, prepare the samples to be loaded
m. Thaw the GeneRuler High Range DNA Ladder and its loading dye at RT
n. Prepare the following mixes to be loaded:

| Sample | Sample amount | Loading Dye | ddH_2_O |
| --- | --- | --- | --- |
| Ladder | 0.5 µl | 1 µl | 4.5 µl |
| PCR reaction | 2.5 µl | 1 µl | 2.5 µl |

o. Move the gel from the cold room (or fridge) into the electrophoresis chamber
p. Make sure the gel is covered with TAE buffer
q. Slowly and carefully remove the comb(s), making sure not to damage the wells
r. Load 6 µl of ladder mix into the correct well(s)
s. Load 6 µl of PCR reaction mix into the correct well(s)
t. Run the gel at 60V for 2-3 hours
u. Carefully place the gel onto the imager and image it

4. **QuBit concentration measurement**
a. Preparation:
i. Equilibrate Qubit dsDNA HS Buffer at RT for 30 min
ii. Thaw enough Qubit dsDNA HS Reagent at RT and keep in a dark place
iii. Prepare enough Qubit tubes and label them, including 2 for the Standards
b. Prepare enough Working Solution, including 10% excess for pipetting errors (make sure to include 2 standards in the calculation):
i. Complete the following table:

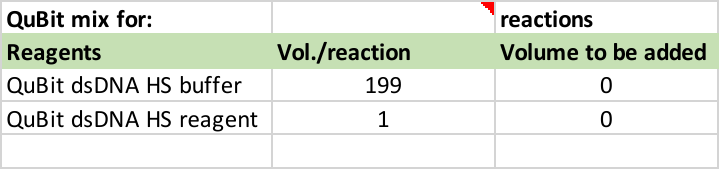

ii. Mix thoroughly by pipetting up and down
c. Add 190 µl of Working solution into the tubes for the Standards
d. Add 199 µl of Working solution into the tubes for the samples
e. Add 10 µl of Standards into the corresponding tubes
f. Add 1 µl of sample into the corresponding tubes
g. Vortex for 3 s
h. Allow all tubes to incubate at RT for 2 min
i. Turn the Qubit fluorometer on
j. Select the dsDNA High Sensitivity measurement
k. Read the Standards
l. Measure the samples and note the results into the Excel sheet:

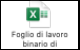

m. Decide how to pool the PCRs together and how the libraries will be composed and fill the following table
   a. Pools can contain all amplicons belonging to the same gDNA template
   b. It is also possible to pool together non-overlapping amplicons from different gDNA template to reduce the number of libraries to be prepared
      i. E.g. amplicons for different loci

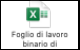

n. Pool all PCR products of the pool together (ideally for at least 130 µl total volume)
5. **DNA shearing**
a. Preparation:
i. If frozen, thaw the PCR pools
ii. Turn Covaris laptop and ultrasonicator on
iii. Fill Covaris well with AFA water to the right level
iv. Place the appropriate Holder XTU
b. Depending on which sequencing reagents will be used, decide on a mean target size for the fragments
i. E.g. to optimise the use of a 500-cycles kit, the ideal mean insert size is about 500-600 bp long
ii. For a 300-cycles kit, the ideal mean insert size is about 300-350 bp long
c. Pipette 130 µl of PCR pool into a 130 µl Covaris AFA microTube
d. Choose an appropriate shearing protocol for the library target size

| Target size (for 130 µl tubes) | Peak incident Power | Duty factor | | Cycles per burst | Treatment Time |
| --- | --- | --- | --- | --- | --- |
| 500 | 50 W | 10% | 200 | | 50 s |
| 400 | 50 W | 10% | 200 | | 70 s |
| 300 | 50 W | 20% | 200 | | 65 s |

e. After shearing, transfer 130 µl of sheared pool into a fresh 1.5-ml tube
6. **Library Quantification and size distribution check with Bioanalyzer**
a. Preparation:
i. Check if DNA High Sensitivity Gel-Dye Mix aliquot is available
   - If not, prepare it as described in the Agilent DNA High Sensitivity protocol
ii. Allow the gel-dye mix to equilibrate at room temperature for 30 min
iii. Allow the marker and ladder to equilibrate at room temperature for 30 min
iv. Assemble priming station
    - Make sure the clip lever is at the lowest position
v. Put a new DNA High Sensitivity chip on the priming station
vi. Make sure the plunger is positioned at 1 ml
b. Pipette 9 µl of gel-dye mix in the well marked **G**
c. Close the priming station
d. Press the plunger until it is held by the clip
e. Wait for exactly 60 s, then release the clip
f. Wait for 5 s and slowly pull the plunger back up at 1 ml
g. Open the priming station
h. Pipette 9 µl of gel-dye mix in the other 2 wells marked with G
i. Pipette 5 µl of marker (green cap) in all 12 sample wells and the ladder well
i. Do not leave any wells empty
j. Pipette 1 µl of DNA ladder (yellow cap) in the well marked with a ladder (bottom right)
k. Pipette 1 µl of sample in the sample wells
l. Pipette 1 µl of water in the unused wells
m. Put the chip horizontally in the chip vortexer and run for 1 min at 2400 rpm
n. Run the chip on the Bioanalyzer within 5 min
o. Fill the table with the mean size of each sample pool

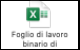

At this point, the sample pools are ready for the NGS library preparation procedure. For this step, different library prep kits are available (e.g. KAPA Hyper, Thruplex DNA-seq, Illumina TruSeq Nano DNA kit). For the TruSeq Nano DNA kit:

7. **Library prep – End Repair**
a. Preparation:
   i. Thaw sheared sample pools at RT
   ii. Equilibrate SPB at RT
   iii. Equilibrate RSB at RT
   iv. Thaw Illumina ERP2 (or ERP3) at RT and then place on ice
   v. Reserve a Thermocycler and make sure the ERP program is saved
b. Distribute 60 µl of sheared sample pools into different 0.2 ml PCR tubes and label them
c. Add 40 µl of ERP2 to each tube
d. Place the PCR tubes/plate on the thermal cycler with the **Illumina TruSeq Nano ERP** program:

| Stage | Temperature (Lid: 100°C) | Time |
| --- | --- | --- |
| End repair | 30°C | 30 min |
| Hold | 4°C | ∞ |

i. Heated lid: 100°C
ii. Total volume: 100 µl
8. **Library prep – Remove large DNA fragments**
a. Preparation:
   i. Prepare an appropriate amount of 80% EtOH (600 µl per 1.5-ml tube) with a 10% excess for pipetting errors
   ii. Vortex SPB until well-dispersed
b. Dilute SPB with ddH_2_O according to the following formula (this formula is for a target size of 350 bp):

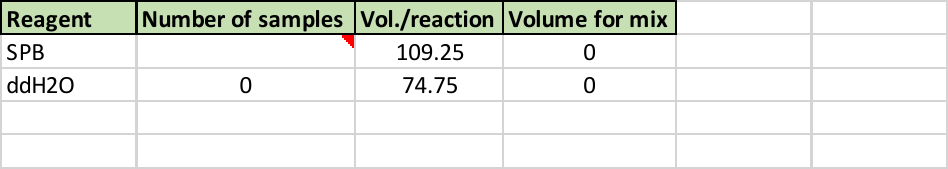

c. Vortex again
d. Transfer each ERP reactions products (100 µl) to a fresh 1.5-ml tube
e. Add 160 µl of diluted SPB to each ERP sample from step b.
f. Vortex and centrifuge briefly
g. Incubate at RT for 5 min
h. Put the tubes onto the appropriate magnet
i. Wait for the solution to clear up
j. After 30 s, rotate the tubes by 90° and repeat 3 more times until they are back to the original position
k. Let stand for an additional 2 min
l. Transfer 250 µl of supernatant to a fresh 1.5-ml tube labeled accordingly
m. Discard remaining diluted SPB
9. **Library prep – Remove small DNA fragments**
a. Preparation:
   i. Prepare an appropriate amount of 80% EtOH (600 µl per 1.5-ml tube) with a 10% excess for pipetting errors
   ii. Vortex SPB until well-dispersed
b. Add 30 µl undiluted SPB to each tube
c. Vortex and centrifuge briefly
d. Incubate at RT for at least 5 min for maximum recovery
e. Put the tubes onto the appropriate magnet
f. Wait for the solution to clear up
g. While keeping the tubes on the magnet, aspirate and discard the supernatant
   i. Leave 5 µl behind to avoid the beads to come with the supernatant
h. Add 300 µl of 80% EtOH to each tube
i. After 30 s, rotate the tubes by 90° and repeat 3 more times until they are back to the original position
j. Remove the supernatant without disturbing the beads
k. Repeat steps h-j
l. Remove residual EtOH with a 20 µl pipette tip
m. Leave the tubes/wells open to dry the residual EtOH for approximately 3.5 min (do not overdry)
n. Remove the tubes from the magnet
o. Add 22 µl of RSB
p. Mix by pipetting up and down
q. Vortex and centrifuge briefly
r. Incubate at RT for 4 min
s. Place the plate/tubes onto the magnet again
t. Wait for the solution to clear up
u. After 30 s, rotate the tubes by 90° and repeat 3 more times until they are back to the original position
v. Transfer 17.5 µl of the eluate to a new PCR tube

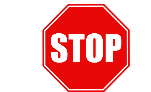

10. **Library prep – A-Tailing**
a. Preparation:
i. Equilibrate RSB at RT for 30 min
ii. Thaw ATL (or ATL2) at RT
iii. Reserve a Thermocycler and make sure the ATAIL70 program is saved
b. Centrifuge ATL (or ATL2) briefly
c. Add 12.5 µl of ATL (or ATL2) to each PCR tube
d. Mix by pipetting up and down
e. Centrifuge briefly
f. Place the PCR tubes/plate on the thermal cycler with the **Illumina TruSeq Nano ATAIL70** program:

| Stage | Temperature (Lid: 100°C) | Time |
| --- | --- | --- |
| A-Tailing | 37°C | 30 min |
| Stop reaction | 70°C | 5 min |
| Cool down | 4°C | 5 min |
| Hold | 4°C | ∞ |

g. Proceed immediately to adapter ligation
11. **Library prep – Adapter ligation**
a. Preparation:
   i. Equilibrate RSB at RT for 30 min
   ii. Equilibrate SPB at RT for 30 min
   iii. Thaw STL at RT
   iv. Thaw selected DNA adapter aliquots at RT
   v. Reserve a thermocycler and make sure the LIG program is saved
b. Briefly centrifuge the adapters
c. Briefly centrifuge the samples
d. Remove LIG2 from the freezer and place on ice
e. Add the following reagents in the order listed to each tube:

| Reagent | Volume |
| --- | --- |
| RSB | 2.5 µl |
| LIG2 | 2.5 µl |
| DNA adapters | 2.5 µl |

f. Mix by pipetting up and down
g. Centrifuge briefly
h. Place the PCR tubes/plate on the thermal cycler with the **Illumina TruSeq Nano LIG** program:

| Stage | Temperature (Lid: 100°C) | Time |
| --- | --- | --- |
| Ligation | 30°C | 10 min |
| Hold | 4°C | ∞ |

i. Volume: 38 µl

i. Centrifuge briefly
j. Immediately add 5 µl of STL to each tube to stop the reaction
k. Transfer 42.5 µl from the PCR tube to a fresh 1.5-ml tube
l. Vortex and centrifuge briefly
12. **Library prep – Clean up ligated fragments**
a. Preparation:
   i. Prepare an appropriate amount of 80% EtOH (600 µl per 1.5-ml tube) with a 10% excess for pipetting errors
   ii. Vortex SPB until well-dispersed
b. Add 42.5 µl undiluted SPB to each tube
c. Vortex and centrifuge briefly
d. Incubate at RT for at least 5 min for maximum recovery
e. Put the tubes onto the appropriate magnet
f. Wait for the solution to clear up
g. While keeping the tubes on the magnet, aspirate and discard the supernatant
   i. Leave 5 µl behind to avoid the beads to come with the supernatant
h. Add 300 µl of 80% EtOH to each tube
i. After 30 s, rotate the tubes by 90° and repeat 3 more times until they are back to the original position
j. Remove the supernatant without disturbing the beads
k. Repeat steps h-j
l. Remove residual EtOH with a 20 µl pipette tip
m. Let the tubes/wells open to dry the residual EtOH for about 3.5 min (do not overdry)
n. Remove the tubes from the magnet
o. Add 54 µl of RSB
p. Mix by pipetting up and down
q. Vortex and centrifuge briefly
r. Incubate at RT for 5 min
s. Place the tubes onto the magnet again
t. After 30 s, rotate the tubes by 90° and repeat 3 more times until they are back to the original position
u. Transfer 50 µl of the eluate to a new 1.5-ml tube
v. Add 50 µl undiluted SPB to each tube
w. Vortex and centrifuge briefly
x. Incubate at RT for at least 5 min for maximum recovery
y. Put the tubes onto the appropriate magnet
z. Wait for the solution to clear up
aa. While keeping the tubes on the magnet, aspirate and discard the supernatant
    i. Leave 5 µl behind to avoid the beads to come with the supernatant
bb. Add 300 µl of 80% EtOH to each tube
cc. After 30 s, rotate the tubes by 90° and repeat 3 more times until they are back to the original position
dd. Remove the supernatant without disturbing the beads
ee. Repeat steps bb-dd
ff. Remove residual EtOH with a 20 µl pipette tip
gg. Leave the tubes open to dry the residual EtOH for approximately 3.5 min (do not overdry)
hh. Remove the tubes from the magnet
ii. Add 29 µl of RSB
jj. Mix by pipetting up and down
kk. Vortex and centrifuge briefly
ll. Incubate at RT for 5 min
mm. Place the tubes onto the magnet again
nn. After 30 s, rotate the tubes by 90° and repeat 3 more times until they are back to the original position
oo. Transfer 25 µl of the eluate to a new PCR tube

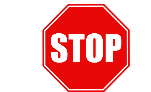

13. **Library prep – Library amplification**
a. Preparation:
i. Equilibrate PPC at RT. Do not vortex
ii. Thaw EPM on ice
iii. If frozen, thaw the samples with beads at RT
iv. Make sure the thermal cycler is reserved and has the amplification program saved
v. If amplifying more than one pooled library, complete reaction mix table on b.
vi. If continuing with purification right away, equilibrate SPB at RT for 30 min
b. Prepare PCR master mix, if necessary:

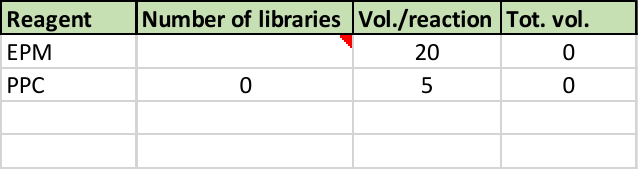

c. If a PCR master mix is prepared, add 25 µl to each tube
d. Otherwise, add 5 µl of PPC and 20 µl of EPM to each tube
e. Mix by pipetting up and down
f. Briefly centrifuge
g. Place the PCR tubes onto the thermal cycler with the **Illumina TruSeq Nano PCR** program:

| Stage | Temperature (Lid: 100°C) | Time | Number of cycles |
| --- | --- | --- | --- |
| Polymerase activation | 95°C | 3 min | 1 |
| Amplification | 98°C | 20 s | 12 |
|  | 60°C | 15 s |  |
|  | 72°C | 30 s |  |
| Final Extension | 72°C | 5 min | 1 |
| Hold | 4°C | ∞ | 1 |

i. Volume: 50 µl
14. **Post-amplification PCR purification**
a. Preparation:
i. Equilibrate SPB at RT for 30 min
ii. Equilibrate RSB at RT for 30 min
iii. Prepare enough fresh 80% ethanol (600 µl per tube)
iv. Vortex the beads until solution is homogeneous
b. Transfer the PCR volume (50 µl) to a fresh 1.5-ml tube
c. After vortexing, add 50 µl (1X volume) of SPB to the amplified libraries
d. Pipette up and down to mix
e. Vortex briefly
f. Incubate at RT for 5 min
g. Put tubes on magnetic stand
h. After 30 s, rotate the tubes by 90° and repeat 3 more times until they are back to the original position
i. With the tube on the magnetic stand, discard the supernatant without touching the beads
   i. Leave 5 µl behind to avoid the beads to come with the supernatant
j. Add 300 µl of 80% ethanol to each tube
k. After 30 s, rotate the tubes by 90° and repeat 3 more times until they are back to the original position
l. Aspirate and discard the supernatant
m. Add 300 µl of 80% ethanol to each tube
n. After 30 s, rotate the tubes by 90° and repeat 3 more times until they are back to the original position
o. Aspirate and discard the supernatant
p. With the tube on the magnetic stand, discard the residual ethanol without touching the beads with a 20 µl tip
q. Dry the beads at RT for 3.5 min
i. Attention: Do not over dry the pellets (over dried pellets appear cracked)
r. Remove tubes from magnet
s. Elute the DNA by re-suspending the beads it in 35 µl of Resuspension Buffer
t. Pipette up and down to mix
u. Centrifuge very briefly
v. Incubate at RT for 5 min
w. Put on magnetic stand
x. After 30 s, rotate the tubes by 90° and repeat 3 more times until they are back to the original position
y. With the tube on the magnetic stand, transfer 31 µl of supernatant into a new tube
ii. If not used immediately, the purified library can be stored at -20°C for up to a week or at 4°C overnight

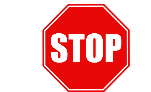

15. **Validate and quantify library**
a. Preparation:
i. Check if High Sensitivity Gel-Dye Mix aliquot is available
   - If not, prepare it as described in the Agilent DNA High Sensitivity Kit protocol
ii. Allow the gel-dye mix to equilibrate at room temperature for 30 min
iii. Allow the marker and ladder to equilibrate at room temperature for 30 min
iv. Assemble priming station
    - Make sure the clip lever is at the lowest position
v. Put a new DNA High Sensitivity chip on the priming station
vi. Make sure the plunger is positioned at 1 ml
b. Pipette 9 µl of gel-dye mix in the well marked **G**
c. Close the priming station
d. Press the plunger until it is held by the clip
e. Wait for exactly 60 s, then release the clip
f. Wait for 5 s and slowly pull the plunger back up at 1 ml
g. Open the priming station
h. Pipette 9 µl of gel-dye mix in the other 3 wells marked with G
i. Pipette 5 µl of marker (green cap) in all 11 sample wells and the ladder well
i. Do not leave any wells empty
j. Pipette 1 µl of DNA ladder (yellow cap) in the well marked with a ladder (position 12)
k. Pipette 1 µl of sample in the sample wells (in duplicate if possible)
l. Pipette 1 µl of water in the not used wells
m. Put the chip horizontally in the chip vortexer and vortex for 1 min at 2400 rpm
n. Run the chip on the Bioanalyzer within 5 min
i. Samples can be stored at -20°C
o. Analyze the size of the libraries and make sure that the mean size increased
   a. Adapters that have been ligated should increase the size of the library
16. **QuBit library concentration measurement**
o. Preparation:
iv. Equilibrate Qubit dsDNA HS Buffer at Rt for 30 min
v. Thaw enough Qubit dsDNA HS Reagent at RT and keep in dark
vi. Prepare enough Qubit tubes and label them, including 2 for the Standards
p. Each library will be measured twice: undiluted and its 1:10 dilution
q. Prepare enough Working Solution, including 10% excess for pipetting errors:
iii. Complete the following table (count each library twice and add the 2 standards)

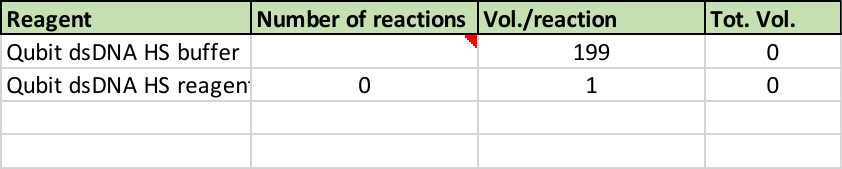

iv. Prepare the master mix accordingly
v. Mix thoroughly by pipetting up and down
r. Add 190 µl of Working solution into the tubes for the Standards
s. Add 199 µl of Working solution into the tubes for the samples
t. Add 10 µl of Standards into the corresponding tubes
u. Add 1 µl of sample into the corresponding tubes
v. Vortex for 3 s
w. Allow all tubes to incubate at RT for 2 min
x. Turn the Qubit fluorometer on
y. Select the dsDNA High Sensitivity measurement
z. Read the Standards
aa. Measure the samples
bb. Annotate the concentration results into the Excel sheet (columns G and H) and add the mean library size from the Bioanalyzer results (column J)

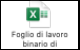

cc. Complete the following Excel sheet to calculate molarity and how to dilute to a target 4 nM

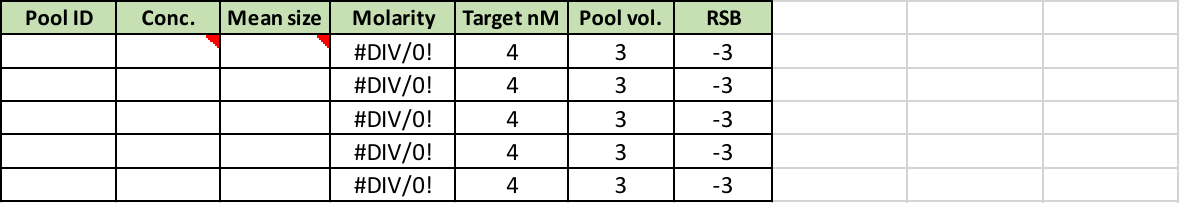

dd. Dilute each library according to column “Pool vol.” and “RSB”
ee. Pool the 4nM libraries into a final 4nM sequencing library proportionally to the total target size of each pool:

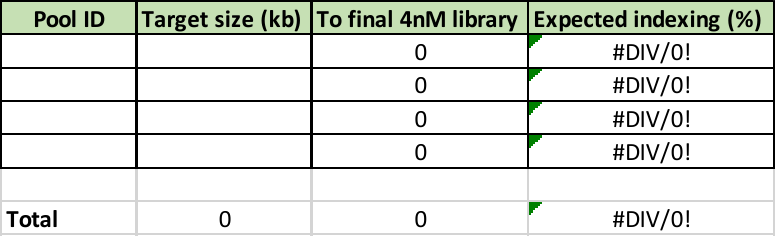

17. **Loading library onto MiSeq**
    a. Preparation:
       i. Thaw 2N NaOH at RT
       ii. Wash the MiSeq instrument, if necessary
       iii. Thaw HT1 at RT
       iv. Chill thawed HT1 on ice (or in the fridge)
       v. Prepare PhiX library, if necessary
       vi. Equilibrate flow cell at RT for 1 hour
       vii. Note the serial number of the reagent cartridge to be used
       viii. Thaw reagent cartridge in water (check max. water level line on the side) for 1 hour
       ix. Place reagent cartridge in fridge until ready to load library
       x. Thaw PR2 bottle in the fridge for 1 hour
    b. Prepare fresh 0.2N NaOH in a fresh 1.5-ml tube:
       i. Add 45 µl of ddH_2_O
       ii. Add 5 µl of 2N NaOH
    c. In a new tube, add 5 µl of final 4 nM library (from step 16 ee)
    d. Add 5 µl of 0.2 N NaOH
    e. Vortex briefly
    f. Centrifuge briefly
    g. Incubate at RT for 5 min
    h. Add 990 µl of pre-chilled HT1 to the denatured library
       i. This results in 1 ml of 20 pM denatured library
       ii. Place on ice
    i. If necessary, dilute to the desired loading concentration:

| Concentration | 8 pM | 10 pM | 12 pM | 15 pM | 20 pM |
| --- | --- | --- | --- | --- | --- |
| 20 pM library | 240 µl | 300 µl | 360 µl | 450 µl | 600 µl |
| Pre-chilled HT1 | 360 µl | 300 µl | 240 µl | 150 µl | 0 µl |

i. Our experience so far tells us that a 12-13 pM library results in a good cluster density
j. Place library ready to be loaded on ice
k. Prepare sample sheet on the instrument with the reagent cartridge serial number information
l. Once the instrument is ready to be used, start the sequencing protocol on it
m. Wash the flow cell carefully to remove solution salts
   i. First spray it with ddH_2_O all over
   ii. Use a lint-free paper tissue to dry it
   iii. Secondly, spray the glass surface on both sides with ethanol, avoiding putting ethanol on the black plastic ports
   iv. Dry and clean the glass surface with a lint-free paper tissue
   v. Once clean and dry, place on instrument according to instruction
n. Follow the instruction from the instrument and load the PR2 bottle and an empty waste bottle
o. Remove reagents cartridge from the fridge
p. Invert at least 10 times to mix
q. Pierce the cartridge well number 17 (highlighted in orange) with a 1000 µl tip
r. Pipette 600 µl of the final denatured library into the well number 17
s. Wait for the software for the prompt to load the reagent cartridge
   i. Remove the wash cartridge
   ii. Insert the reagent cartridge **containing the library** until it cannot slide anymore
t. Close the cartridge door
u. Click next and check if the correct sample sheet will be loaded and if the sequencing details are correct
v. Press next
   i. At this point the instrument will check its functionality and the fluidic systems
w. Once the pre-run check is done, click on Start
x. The run will start
y. The cluster density info will appear after cycle 15 (roughly 2-3 hours)
z. The Q30 statistics will appear after cycle 25 (roughly 3h 30 min)
